# Germline *RUNX1* Variation and Predisposition to Childhood Acute Lymphoblastic Leukemia

**DOI:** 10.1101/2021.04.29.21256340

**Authors:** Yizhen Li, Meenakshi Devidas, Wentao Yang, Stuart S. Winter, Wenjian Yang, Kimberly P. Dunsmore, Colton Smith, Maoxiang Qian, Xujie Zhao, Ranran Zhang, Julie M. Gastier-Foster, Elizabeth A. Raetz, William L. Carroll, Chunliang Li, Paul P. Liu, Karen R. Rabin, Takaomi Sanda, Charles G. Mullighan, Kim E. Nichols, William E. Evans, Ching-Hon Pui, Stephen P. Hunger, David T. Teachey, Mary V. Relling, Mignon L. Loh, Jun J. Yang

## Abstract

RUNX1 is a transcription factor critical for definitive hematopoiesis and genetic alterations in *RUNX1* have been implicated in both benign and malignant blood disorders, particularly of the megakaryocyte and myeloid lineages. Somatic *RUNX1* mutations are reported in B- and T-cell acute lymphoblastic leukemia (B-ALL and T-ALL), but germline genetic variation of *RUNX1* in these lymphoid malignancies have not been comprehensively investigated. Sequencing 4,836 children with B-ALL and 1,354 cases of T-ALL, we identified 31 and 18 unique germline *RUNX1* variants in these two ALL subtypes, respectively. *RUNX1* variants in B-ALL were predicted to have minimal impact. By contrast, 54.5% of variants in T-ALL result in complete or partial loss of RUNX1 activity as a transcription activator *in vitro*, with dominant negative effects for 4 variants. Ectopic expression of dominant negative deleterious *RUNX1* variants in human CD34+ cells repressed differentiation into erythroid, megakaryocytes, and T cells, while promoting differentiation towards myeloid cells. We then performed chromatin immunoprecipitation profiling in isogenic T-ALL models with variants introduced by genome editing of endogenous *RUNX1*. We observed highly distinctive patterns of DNA binding and target genomic loci by RUNX1 proteins encoded by the truncating vs missense variants. The p.G365R *RUNX1* variant resulted in a novel methylation site in RUNX1 and alteration in its interaction with CBFβ. Further whole genome sequencing showed that *JAK3* mutation was the most frequent somatic genomic abnormality in T-ALL with germline *RUNX1* variants. Consistently, co-introduction of *RUNX1* variant and *JAK3* mutation in hematopoietic stem and progenitor cells in mouse gave rise to T-ALL with early T-cell precursor phenotype *in vivo*, compared to thymic T-ALL seen in mice with *JAK3* mutation alone. Taken together, these results indicated that *RUNX1* is an important predisposition gene for ALL, especially in T-ALL and also pointed to novel biology of RUNX1-mediated leukemogenesis in the lymphoid lineages.

## Introduction

Acute lymphoid leukemia (ALL) is the most common cancer in children. The exact cause of ALL is incompletely understood, although somatic genomic abnormalities are well documented affecting a wide range of signaling pathways. There is also growing evidence of inherited susceptibility to ALL. For example, common genetic polymorphisms in genes such as *IKZF1*(1), *ARID5B* (2), *CDKN2A*(3), *GATA3* (4, 5), *CEBPE*(6), and *PIP4K2A*(7) are associated with the risk of ALL in an age- and subtype-dependent manner. On the other hand, rare germline variants have been linked to familial predisposition to childhood ALL, and collectively about 5% of sporadic ALL cases harbor pathogenic variants in *TP53*(8), *ETV6*(9), and *IKZF1*(1). These findings point to a strong genetic basis of inter-individual variability in ALL risk.

The RUNX1 protein plays key roles in definitive hematopoiesis (10). RUNX1 functions as a transcription factor by forming a heterodimer with core binding factor β (CBFβ). RUNX1 consists of a Runt homology domain (RHD) responsible for DNA binding and cofactor interaction (11) and the C-terminal transcriptional activation domain (TAD) that recruits co-activators and activates the expression of RUNX1 target genes (12). *RUNX1* germline variants are associated with familial platelet disorder (FPD). Many patients with FPD develop leukemia later in life, predominately acute myeloid leukemia (AML) and myelodysplastic syndrome (MDS) (13-16). Somatic *RUNX1* mutations, most of which occur in the RHD and TAD, have been identified in both B- and T-ALL(17). *RUNX1* mutation is related to poor prognosis in T-ALL(17). Although somatic and germline *RUNX1* variants associated with ALL have been reported, their pattern, prevalence, and functional consequences in B-ALL and T-ALL have not been comprehensively investigated.

Here we report results from targeted germline sequencing of 6,190 children with B- or T-ALL enrolled in frontline Children’s Oncology Group (COG) and St. Jude Children’s Research Hospital (St. Jude) ALL trials. We observed a lineage-specific pattern of germline variation in the *RUNX1* gene, with deleterious variants exclusively present in T-ALL patients. Furthermore, we experimentally characterized *RUNX1* variants for their effects on transcription factor activity, subcellular localization, cofactor interaction, *in vitro* hematopoiesis, and genome wide RUNX1 binding profile. Finally, we examined the somatic genomic landscape of T-ALL arising from *RUNX1* germline variants and modeled *RUNX1*-mediated leukemogenesis in mouse models.

## Results

### Identification of germline *RUNX1* variants in pediatric ALL

To comprehensively characterize inherited *RUNX1* variations in ALL, we performed targeted sequencing in germline DNA of 4,836 patients with newly diagnosed B-ALL and 1,354 patients with T-ALL enrolled on COG and SJCRH frontline trials (**Figure 1A and Table 1**). We identified 31 unique variants in 61 B-ALL cases and 18 unique variants in 26 T-ALL cases. Seven of these variants were found in both B- and T-ALL (**Figure 1A and Table 1**).

**Figure 1.**
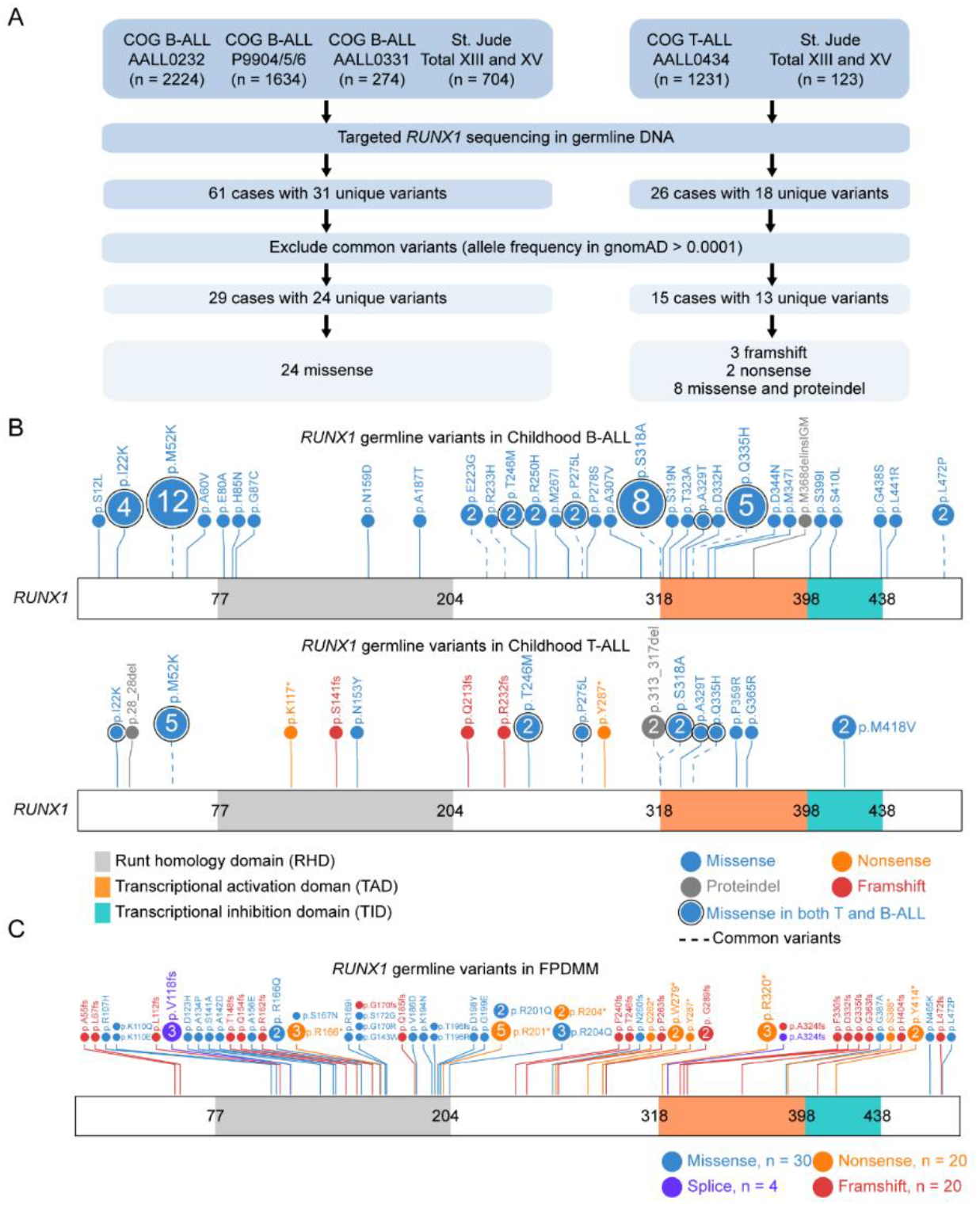
Germline *RUNX1* variants in childhood B- and T-ALL. (A) CONSORT diagram of the Children’s Oncology Group (COG) and St. Jude Children’s Research Hospital (St. Jude) patients included in this study. (B) Protein domain plot of RUNX1 and the amino acid substitutions predicted to result from the germline *RUNX1* variants identified in this study. The upper panel showed germline *RUNX1* variants in B-ALL cases, and the lower panel showed those in T-ALL cases. (C) Protein domain plot of RUNX1 and the germline RUNX1 variants identified previously in familial platelet disorder with associated myeloid malignancy (FPDMM). Data were retrieved from recently published paper (18).

**Table 1.** Germline RUNX1 variants in pediatric ALL cases.

| Protein (NM 001754) | CHROM | POS | REF | ALT | ExonicFunc.refGene | ALL Subtype (n = case number) | Allele frequency in gnomAD | Polyphen2_HDIV_pred | SIFT_pred |
| --- | --- | --- | --- | --- | --- | --- | --- | --- | --- |
| p.L472P | 21 | 36164460 | A | G | missense | B-ALL (2) | 1.99E-04 | Probably damaging | Deleterious |
| p.L441R | 21 | 36164553 | A | C | missense | B-ALL (1) | 7.28E-06 | Probably damaging | Deleterious |
| p.G438S | 21 | 36164563 | C | T | missense | B-ALL (1) |  | Probably damaging | Tolerated |
| p.S410L | 21 | 36164646 | G | A | missense | B-ALL (1) | 1.56E-05 | Probably damaging | Deleterious |
| p.S399I | 21 | 36164679 | C | A | missense | B-ALL (1) | 1.74E-05 | Probably damaging | Deleterious |
| p.M368delinsIGM | 21 | 36164771 | C | >ATGCC | nonframeshift_insertion | B-ALL (1) | 7.34E-05 | . | . |
| p.M347I | 21 | 36164834 | C | T | missense | B-ALL (1) | 8.79E-06 | Benign | Deleterious |
| p.D344N | 21 | 36164845 | C | T | missense | B-ALL (1) | 6.20E-05 | Probably damaging | Tolerated |
| p.Q335H | 21 | 36164870 | C | A | missense | B-ALL (5) and T-ALL (1) | 1.70E-04 | Probably damaging | Deleterious |
| p.D332H | 21 | 36164881 | C | G | missense | B-ALL (1) | 4.73E-06 | Probably damaging | Deleterious |
| p.A329T | 21 | 36164890 | C | T | missense | B-ALL (1) and T-ALL (1) |  | Possibly damaging | Tolerated |
| p.T323A | 21 | 36171598 | T | C | missense | B-ALL (1) | 1.59E-05 | Benign | . |
| p.S319N | 21 | 36171609 | C | T | missense | B-ALL (1) |  | Possibly damaging | Tolerated |
| p.S318A | 21 | 36171613 | A | C | missense | B-ALL (8) and T-ALL (2) | 8.13E-04 | Possibly damaging | Tolerated |
| p.A307V | 21 | 36171645 | G | A | missense | B-ALL (1) |  | Possibly damaging | Tolerated |
| p.P278S | 21 | 36171733 | G | A | missense | B-ALL (1) | 1.19E-05 | Possibly damaging | Tolerated |
| p.P275L | 21 | 36171741 | G | A | missense | B-ALL (2) and T-ALL (1) | 2.03E-04 | Possibly damaging | Tolerated |
| p.M267I | 21 | 36206711 | C | T | missense | B-ALL (1) | 3.93E-05 | Benign | Tolerated |
| p.R250H | 21 | 36206763 | C | T | missense | B-ALL (2) | 3.91E-05 | Probably damaging | Tolerated |
| p.T246M | 21 | 36206775 | G | A | missense | B-ALL (2) and T-ALL (2) | 4.61E-05 | Probably damaging | Tolerated |
| p.R233H | 21 | 36206814 | C | T | missense | B-ALL (1) | 1.67E-04 | Probably damaging | Tolerated |
| p.E223G | 21 | 36206844 | T | C | missense | B-ALL (2) | 1.24E-04 | Probably damaging | Deleterious |
| p.A187T | 21 | 36231825 | C | T | missense | B-ALL (1) | 3.98E-06 | Probably damaging | Deleterious |
| p.N159D | 21 | 36252887 | T | C | missense | B-ALL (1) |  | Probably damaging | Deleterious |
| p.G87C | 21 | 36259232 | C | A | missense | B-ALL (1) |  | Probably damaging | Deleterious |
| p.H85N | 21 | 36259238 | G | T | missense | B-ALL (1) | 3.59E-05 | Probably damaging | Deleterious |
| p.E80A | 21 | 36259252 | T | G | missense | B-ALL (1) | 1.22E-05 | Possibly damaging | Deleterious |
| p.A60V | 21 | 36259312 | G | A | missense | B-ALL (1) | 7.40E-05 | Possibly damaging | Tolerated |
| p.M52K | 21 | 36259336 | A | T | missense | B-ALL (12) and T-ALL (5) | 2.46E-04 | Probably damaging | Deleterious |
| p.I22K | 21 | 36265254 | A | T | missense | B-ALL (4) and T-ALL (1) | 4.01E-06 | Benign | Tolerated |
| p.S12L | 21 | 36421162 | G | A | missense | B-ALL (1) | 3.98E-06 | Benign | Tolerated |
| p.M418V | 21 | 36164623 | T | C | missense | T-ALL (2) |  | Benign | Deleterious |
| p.G365R | 21 | 36164782 | C | G | missense | T-ALL (1) | 9.73E-06 | Probably damaging | Deleterious |
| p.P359R | 21 | 36164799 | G | C | missense | T-ALL (1) |  | Probably damaging | Deleterious |
| p.313_317del | 21 | 36171614 | TTCTGCA | A | nonframeshift_deletion | T-ALL (2) | 1.73E-04 | . | . |
| p.Y287X | 21 | 36171704 | G | C | stopgain | T-ALL (1) |  | . | Tolerated |
| p.R232fs | 21 | 36206815 | GC | G | frameshift_deletion | T-ALL (1) |  | . | . |
| p.Q213fs | 21 | 36206874 | TG | T | frameshift_deletion | T-ALL (1) |  | . | . |
| p.N153Y | 21 | 36252905 | T | A | missense | T-ALL (1) |  | Probably damaging | Deleterious |
| p.S141fs | 21 | 36252939 | C | CGGTT | frameshift_insertion | T-ALL (1) | 2.39E-05 | . | . |
| p.K117X | 21 | 36259142 | T | A | stopgain | T-ALL (1) |  | . | Tolerated |
| p.28_28del | 21 | 36265234 | TAGA | T | nonframeshift_deletion | T-ALL (1) | 4.62E-05 | . | . |

Polyphen2, Polymorphism Phenotyping v2; SIFT, Sorting Intolerant From Tolerant.

Of the 31 variants in B-ALL, 6 were not observed in the general population (Genome Aggregation database, gnomAD, n = 15,496), 18 were rare with a maximum allele frequency of 0.00122%, and the remaining 7 were considered common variants with allele frequency > 0.01% (**Figure 1A and Table 1**). All the variants in B-ALL except one were missense, most of which are in the C-terminus distal to the DNA binding runt-homology domain (RHD, **Figure 1B**). Of the 18 variants in T-ALL cases, 8 were absent in the gnomAD dataset, 5 were rare with a maximum allele frequency of 0.00239%, and the remaining 5 were common variants (**Figure 1A and Table 1**). 27.8% of variants identified in T-ALL were frameshift or nonsense, including p.K117* and p.S141fs which truncated both the RHD and the transcription activation domain (TAD, **Figures 1B and S1**) and p.Q213fs, p.R232fs, and p.Y287* that resulted in the loss of TAD only (**Figure S1**). Seven missense and 1 in-frame deletion variants in T-ALL were distributed across *RUNX1*. This pattern of variant distribution is significantly different from *RUNX1* germline variants in familial platelet disorder with associated myeloid malignancy (**Figure 1C**)(18), in which the majority of missense variants are localized in the DNA-binding domain (RHD).

### Effects of *RUNX1* variants on transcriptional regulation, cellular localization, and protein– protein interaction

To understand how germline *RUNX1* variants affect gene function, we first examined their transcription activator activity using the luciferase reporter assay in Hela cells. With *SPI1* as the RUNX1 target gene (19), none of the germline variants identified in B-ALL showed a significant impact on reporter gene transcription compared to the wildtype protein and therefore were not studied further (**Figure 2A**). Among *RUNX1* alleles seen in T-ALL, all frameshift and nonsense variants (p.K117*, p.S141fs, p.S213fs, p.R232fs, and p.Y287*) and also missense variant G365R caused significant reduction of RUNX1 activity in this assay (**Figure 2B**). To further characterize these *RUNX1* variants in a more relevant cellular context, we engineered the Jurkat T-ALL cell line in which each *RUNX1* variant of interest was individually inserted into the safe harbor AAVS1 locus (**Figure S2**)(20) and a GFP tag was added to the C-terminus of RUNX1 target gene *GZMA(21)* (**Figure 2C and S3**). Using this model system, RUNX1 transactivation activity could be directly measured as the GFP intensity in *RUNX1* variants knock-in cells, in the presence of endogenous *RUNX1* (**Figure 2D**). As shown in **Figure 2E**, the introduction of WT *RUNX1* as well as most missense variants (p.N153Y, p.T246M, p.A329T, p.P359R, and p.M418V) led to robust GFP signals relative to cells with no *RUNX1* insertion at the AAVS locus, confirming wildtype like transcription activator activity. By contrast, cells with K117* and S141fs showed only baseline GFP signals, indicating complete loss of RUNX1 function. Insertion of the p.Q213fs, p.R232fs, p.Y287*, and p.G365R variants resulted in the lowest GFP intensity, suggesting these variants not only lost their transcription activator activity but also repressed endogenous RUNX1 in a plausibly dominant-negative manner.

**Figure 2.**
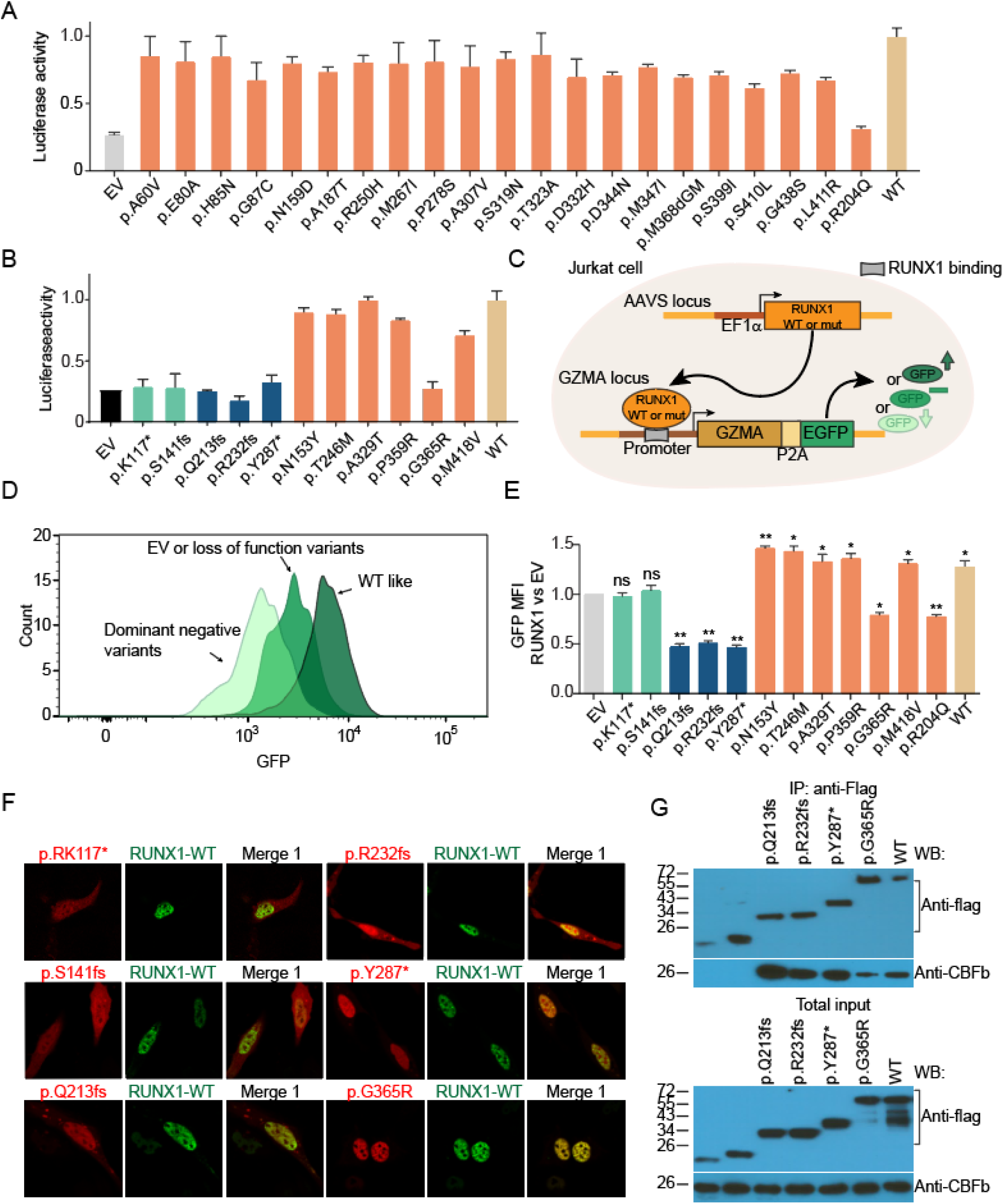
Germline *RUNX1* variants influence transcription factor activity, subcellular localization, and CBFβ interaction. (A) Luciferase reporter gene assay (driven by the *PU*.*1* promoter in Hela cells) showed minimal effects on transcription factor activity by missense *RUNX1* variants identified in B-ALL. (B) By contrast, 6 of 11 *RUNX1* variants observed in T-ALL resulted in complete or partial loss of activity, as measured using luciferase reporter gene assay. (C) Design of the Jurkat landing-pad system to measure RUNX1 variant activity in T-ALL. *RUNX1* (either WT or variant) was inserted at the AAVS locus. EGFP coding sequence was knocked at the 3’-end of *GZMA*, a RUNX1 target gene. RUNX1 transcription factor activity was determined by flow cytometry of GFP signal which reflects RUNX1-driven *GZMA* transcription. (D) Flow cytometry analysis of Jurkat cells expressing different *RUNX1* variants. Cells harboring dominant negative, loss of function, and WT like RUNX1 variants exhibited the lowest, moderate, and highest GFP signals, respectively. (E) The GFP signal from Jurkat cells expressing each *RUNX1* variant (relative to empty vector) is shown in bar graph, with error bars indicating standard deviation of triplicates. (F) Immunofluorescence microscopy shows subcellular localization of mCherry-tagged variant proteins and EGFP-tagged WT RUNX1. Variant and WT RUNX1 were fused to mCherry and EGFP and expressed transiently in HEK293T cells, which were then subjected to imaging analyses. (G) Co-immunoprecipitation assay was performed to determine RUNX1-CBFβ interaction for each deleterious variant. Experiments were performed in HEK293T cells. RUNX1 proteins were pulled down using anti-FLAG antibody and the presence or absence of CBFβ in the pellet was examined by immunoblotting.

We next analyzed subcellular localization and CBFβ cofactor interaction of all deleterious variants, including p.K117*, p.S141fs, p.Q213fs, p.R232fs, p.Y287*, and p.G365R (**Figure S1**). Fluorescence microscopy of HEK293T cells ectopically expressing *RUNX1* variants showed that p.K117* and p.S141fs proteins were mis-localized to the cytoplasm, whereas p.Q213fs, p.R232fs, p.Y287*, p.G365R proteins remained in the nucleus (**Figure 2F**). In co-immunoprecipitation assay, p.K117* and p.S141fs variant proteins were no longer associated with CBFβ most likely due to the absence of RHD, whereas all the other variants retained the ability to interact with this co-factor (**Figure 2G**).

### Effects of *RUNX1* variants on the differentiation and proliferation of human cord blood CD34+ cells *in vitro*

We next sought to examine the effects of *RUNX1* variants on hematopoietic differentiation *in vitro* using human cord blood CD34+ cell as the model system. Because p.K117* and p.S141fs simply resulted in complete loss of function with no dominant negative effects, we chose not to further characterize them. For the remaining deleterious variants, we selected p.R232fs, p.Y287* and p.G365R to represent frameshift, nonsense, and missense variants, respectively (**Figure S4**). RUNX1 variants were ectopically expressed in human CD34+ cells which were then subjected to differentiation, proliferation, and apoptosis assays *in vitro* (**Figure 3A**).

**Figure 3.**
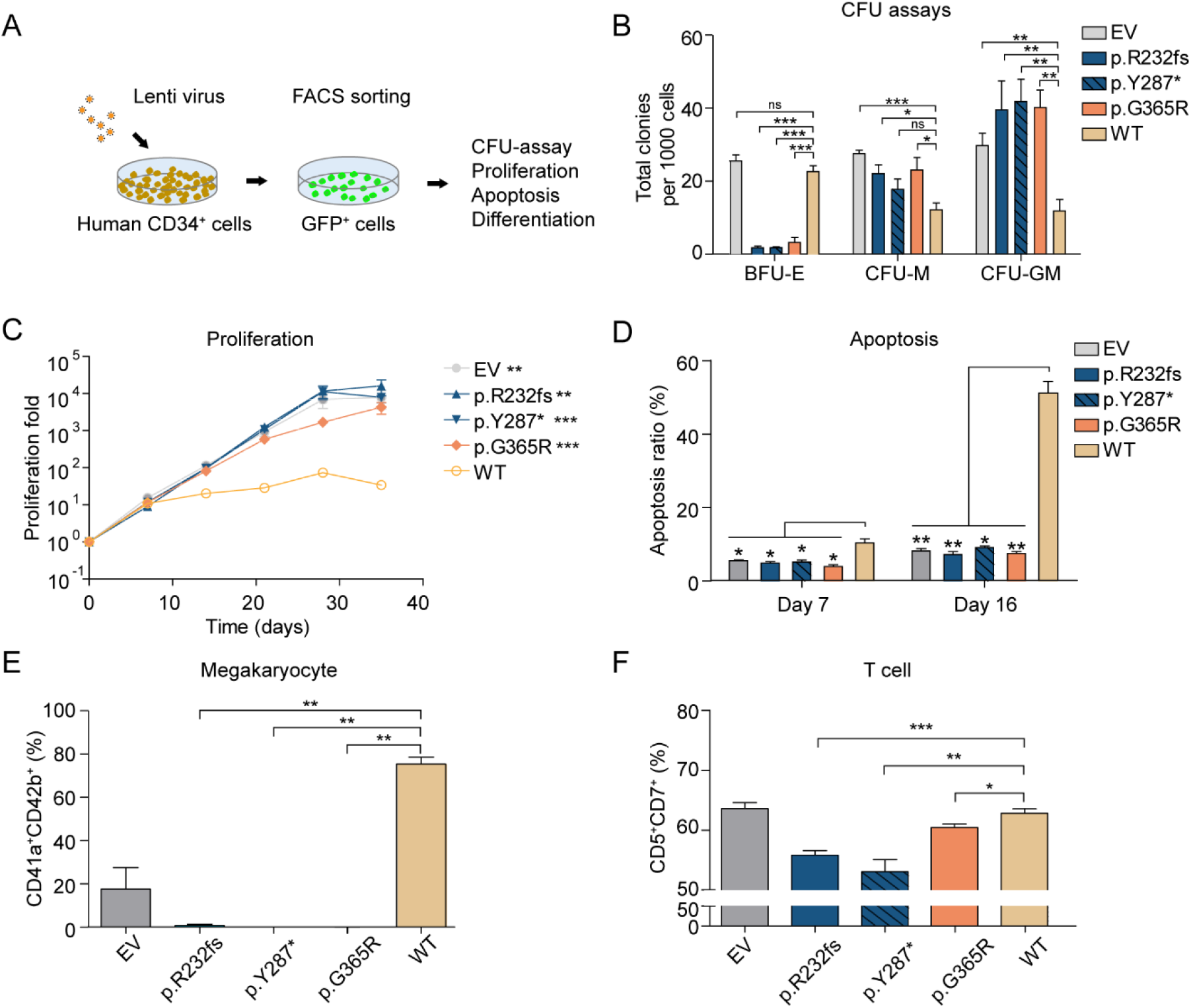
*RUNX1* variants affect *in vitro* differentiation of human cord blood CD34+ cells. (A) The scheme shows the design of in vitro hematopoietic differentiation assay. *RUNX1* variants were lentivirally introduced into human cord blood CD34+ cells. Successfully transduced cells were sorted by flow cytometry and processed for colony forming unit assays, and assessed for cell proliferation and apoptosis, as appropriate. (B) One thousand RUNX1-expressing CD34+ cells were plated in MethoCult H4034. The Y-axis shows the count of colonies for each lineage: burst-forming unit erythroid (BFU-E), colony-forming unit-macrophage (CFU-M), and colony-forming unit granulocyte-macrophage (CFU-GM). (C) Proliferation of RUNX1-expressing CD34+ cells were monitored for 5 weeks, in Iscove’s Modified Dulbecco Medium (IMDM) medium containing 20% BIT9500, 10 ng/mL FLT-3 ligand, TPO, SCF, IL-3, and IL-6. The number of cells was counted every week for 5 weeks. (D) Apoptosis of RUNX1-transduced CD34+ cells after 7 and 16 days of culture (same culture medium as C) was measured by flow cytometry using Annexin-V and DAPI antibodies. (E-F) CD34+ cells ectopically expressing *RUNX1* variants were also subjected to *in vitro* differentiation assays for megakaryocyte or T cell lineages. Following RUNX1 transduction, cells were cultured in the presence of SFEMII containing megakaryocyte expansion supplement or T-Cell progenitor differentiation supplement for 2 weeks. Megakaryocyte (E) was identified as CD41a+/CD42b+, and T cells (F) were defined as CD5+/CD7+ by flow cytometry.

In colony formation assays conditioned for erythroid and myeloid progenitor cell growth, the expression of p.R232fs, p.Y287*, and p.G365R significantly repressed burst-forming unit erythroid (BFU-E) and increased colony-forming unit granulocyte-macrophage (CFU-GM) colonies compared to CD34+ cells transduced with WT RUNX1 (**Figure 3B**). The immunophenotype of these progenitor cells were also confirmed by flow cytometry (**Figure S5A**). Long-term culture showed that the *RUNX1* variant-transduced CD34+ cells proliferated faster with concomitant reduction in apoptosis, compared to WT *RUNX1* transduced cells (**Figure 3C-D and S5B**).

With culture conditions for megakaryocyte differentiation, expression of RUNX1 variants consistently resulted in a significant reduction of CD41a+/CD42b+ population compared to WT (**Figure 3E**). These RUNX1 variants also significantly repressed the generation of CD5+/CD7+ T cells from the CD34+ population (**Figure 3F**). Collectively, these results suggested that *RUNX1* variants promoted myeloid differentiation while repressing megakaryocyte and T-cell differentiation *in vitro*.

### *RUNX1* variants have highly distinctive patterns of DNA binding and are associated with altered post-translational modifications

To understand the molecular effects of *RUNX1* variants, we comprehensively profiled RUNX1 binding across the genome using chromatin immunoprecipitation-sequencing (ChIP-seq). We first engineered three isogenic Jurkat cell lines in which each of the three *RUNX1* variants (p.R232fs, p.Y287*, and p.G365R) was individually knocked-in at the endogenous locus in a hemizygous fashion to represent heterozygous genotype seen in patients (**Figure 4A and 4B**). In these models, we introduced the HA tag and TY1 epitope tags at the 3’ end of the coding exon on the variant and WT *RUNX1* alleles, respectively (**Figure S6-S10**). This enabled us to separately profile variant or WT RUNX1 binding using HA or TY1 antibodies (**Figure S9C-D and S10C-D**). We also generate two single clones, in which both allele were wildtype *RUNX1*, but tagged with HA and TY1 seperately (**Figure S8 and S10**).

**Figure 4.**
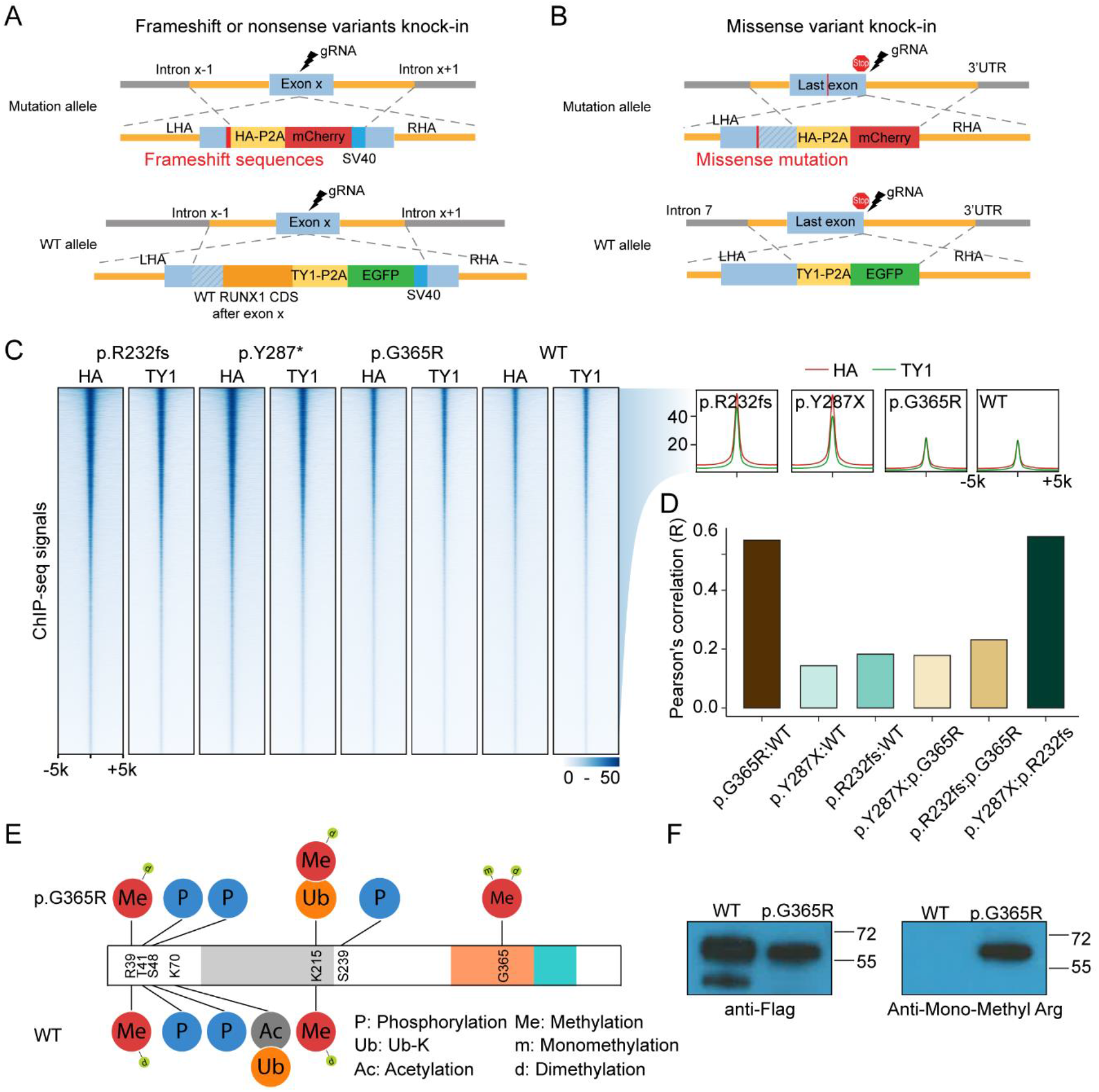
*RUNX1* variants have highly distinctive DNA binding patterns and associated with altered post-translational modifications. (A-B) Schematic representation of engineered Jurkat cell models for RUNX1 binding profiling studies. Variants (p.R232fs, p.Y287*, and p.G365R) were knocked in using CRISPR-cas9 editing at the endogenous locus in a heterozygous fashion. Meanwhile TY1 and HA epitopes were inserted to the coding sequence of WT and variant RUNX1, respectively. This design enables ChIP-seq of each protein simultaneously using two different antibodies. (C) ChIP-seq using HA antibody (recognizing RUNX1 variant) or TY1 antibody (for wildtype RUNX1) showed that activation domain truncating variant RUNX1 (p.R232fs and p.Y287*) recognized largely identical genomic regions as missense variant p.G365R and wildtype RUNX1. Cells with homozygous WT genotype were used as control. (D) Pearson’s correlation of WT and variant RUNX1 ChIP-seq signals (HA divided by TY1). (E) Predicted post translational modification of the p.G365R protein showed arginine methylation resulting from the single nucleotide variant. (F) Arginine mono-methylation was confirmed by immunoblotting using an anti-Mono-Methyl Arginine antibody.

ChIP-seq showed that all three variants have a largely overlapping binding profile as WT RUNX1 in T-ALL genome (**Figure 4C**). However, the C-terminal truncating variants p.R232fs and p.Y287* exhibited a much similar binding pattern compared with missense p.G365R variant and wildtype RUNX1 (**Figure 4D and S11**), as evidenced by the pearson correlation coefficient of ChIP-seq signals. Even though these variant proteins maintained the DNA binding domain, their DNA binding preference was different from wildtype or full-length missense mutation RUNX1.

Interestingly, the p.G365R variant gave rise to a novel methylation site in RUNX1, with mono- or di-methylation of the arginine residue confirmed by mass spectrometry and Western blot analysis (**Figure 4E and 4F**). Immunoprecipitation–mass spectrometry results suggested that RUNX1 protein methylation at this site may disrupt its interaction with TUBB family proteins (TUBB2A, TUBB2B, TUBB4B, TUBB5, TUB8, *et al*.) and heat shock proteins, but with an increase of CBFβ binding (**Table S1**).

### Somatic genomic abnormalities in T-ALL with germline *RUNX1* variants

To characterize the somatic genomic landscape of T-ALL with germline *RUNX1* variants, we analyzed whole genome seq of six cases with p.K117*, p.S141fs, p.Q213fs, p.R232fs, p.Y287*, and p.G365R variants, which were contrasted with 263 T-ALL with somatic mutations in *RUNX1* or WT genotype (22). Five of six T-ALL (83.3%) with germline *RUNX1* variants had a somatic *JAK3* mutation, significantly higher compared to the frequency of *JAK3* mutation percentage in T-ALL cases without germline variants in *RUNX1* (7.6%, p-value = 2.59×10^−5^) (22) or T-ALL cases with somatic mutations in *RUNX1* (27.3%, p-value = 0.05) (**Figure 5A**). *JAK3* mutations in T-ALL cases with germline *RUNX1* variants were located in either the pseudo-kinase domain (M511I and R657Q) or in the kinase domain (L950V, **Figure S12 and Tables S2 and S3**). Of interest, the patient with a germline *RUNX1-*R232fs variant also subsequently acquired a somatic *RUNX1* mutation (R169_E5splice_region).

**Figure 5.**
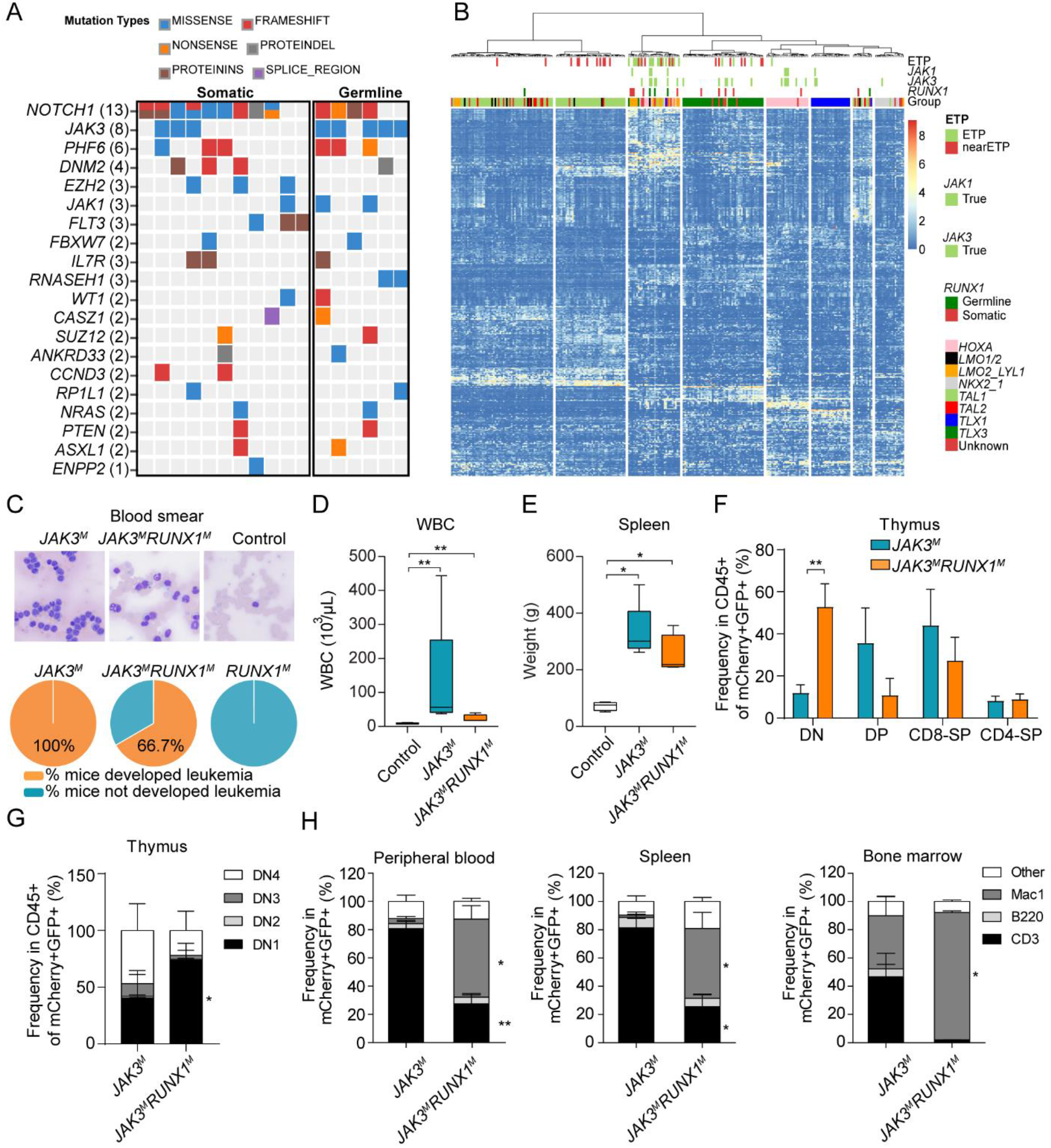
Somatic *JAK3* mutations co-occurs in T-ALL with germline *RUNX1* variants and jointly drive ETP phenotype in mouse models. (A) Somatic *JAK3* mutations were significantly enriched in T-ALL cases with germline *RUNX1* variants. Whole genome seq of remission samples for 17 T-ALL cases, 6 and 11 with germline variants or somatic mutations in *RUNX1*, respectively. (B) RNA-seq was analyzed for 267 T-ALL cases, including 252, 4, and 11 subjects with WT RUNX1, carrying germline variants or somatic mutations in this gene. Unsupervised clustering shows that RUNX1-variant cases, either germline or somatic, clustered tightly with T-ALL with ETP and near-ETP immunophenotypes. (C) Upper panel: examples of blood smear of *JAK3*^*M*^ and *JAK3*^*M*^*RUNX1*^*M*^ mice at the time of sacrifice, and control mice after 4 months of transplantation. Lower panel: the percentage of mice developed leukemia in each group. *JAK3*^*M*^: 100%, 5 out of 5; *JAK3*^*M*^*RUNX1*^*M*^: 66.7%, 4 out of 6. (D) Peripheral leukocyte count of *JAK3*^*M*^*RUNX1*^*M*^ (n = 4) and *JAK3*^*M*^ (n = 5) at the time of sacrifice, and control (n = 7) mice after 4 months of transplantation. (E) Spleen weight of *JAK3*^*M*^*RUNX1*^*M*^ (n = 4), and *JAK3*^*M*^ (n = 5) mice at the time of sacrifice and control mice (n = 4) after 4 months of transplantation. (F and G) Thymocyte immunophenotype of *JAK3*^*M*^ and *JAK3*^*M*^*RUNX1*^*M*^ mice at the time of sacrifice. Co-expression of *RUNX1*^*M*^ and *JAK3*^*M*^ resulted in a drastic increase in DN1 population compared with mice receiving LSK cells expressing JAK3^M^ only. (H) In peripheral blood, bone marrow, and spleen, *JAK3*^*M*^*RUNX1*^*M*^ mice showed a significant increase in Mac1+ population and a reduction of the CD3+ population compared to *JAK3*^*M*^ mice.

We also performed RNA-seq of T-ALL with germline *RUNX1* variants and compared the expression profile with cases with germline *RUNX1* variants, somatic *RUNX1* mutations or WT *RUNX1* (N = 4, 11 and 252, respectively). Based on hierarchical clustering of global expression profile, *RUNX1*-variant cases (either germline or somatic) consistently clustered with T-ALL with early T-cell precursor immunophenotype (ETP) or near-ETP cases (**Figure 5B**). These results are consistent with previous reports of the preponderance of *RUNX1* variants in ETP T-ALL (23).

### *RUNX1* and *JAK3* mutation-induced ETP phenotype in murine bone marrow transplantation model

To model *RUNX1*-related T-ALL leukemogenesis, especially in conjunction with somatic *JAK3* mutation, we introduced different combinations of *RUNX1* and *JAK3* mutations (*RUNX1*^*R232fs*^ and *JAK3*^*M511I*^) into mouse hematopoietic progenitor cells (Lin^−^/Sca-1^+^/C-Kit^+^) and monitored leukemia development *in vivo* after transplantation. We hereafter refer to recipient mice with LSK cells transduced with empty vector, *RUNX1*^*R232fs*^, *JAK3*^*M511I*^, and *JAK3*^*M511I*^*/RUNX1*^*R232fs*^ as “control”, “*RUNX1*^*M*^”, “*JAK3*^*M*^”, and “*JAK3*^*M*^*RUNX1*^*M*^” mice, respectively. At 4 months, peripheral leukocyte counts of *JAK3*^M^ and *JAK3*^*M*^*RUNX1*^*M*^ mice (41.78 ± 44.3 E3 cells/µL and 14.93 ± 3.42 E3 cells/µL respectively) were significantly higher than control mice (8.84 ± 2.00 cells/µL), and the lowest peripheral leukocyte counts were seen in *RUNX1*^*M*^ mice (6.10 ± 2.03 cells/µL, **Figure S13A**). Flow cytometry analysis at this time point showed a significant increase of CD8+ T cells in *JAK3*^M^ mice (**Figure S13B and C**), compared with control mice. By contrast, *JAK3*^M^*RUNX1*^M^ mice showed an increase in Mac1+ population and lower T cell population, suggesting an outgrowth of cells with ETP immunophenotype (**Figure S13B and C**).

At 6 to 10 months after transplantation, both *JAK3*^M^*RUNX1*^M^ and *JAK3*^M^ mice developed overt leukemia presented with leukocytosis and splenomegaly, with 66.7% and 100% mice developed leukemia, respectively (**Figure 5C-E and S14**). The thymus of *JAK3*^M^*RUNX1*^M^ mice showed a significantly increase of CD4-CD8-(DN) T cells, particularly DN1 cells, as compared with *JAK3*^M^ mice (**Figure 5F-G**). Circulating leukemic cells of *JAK3*^M^*RUNX1*^M^ mice showed a markedly higher Mac1+ population, but lower lymphoid surface marker as compared with *JAK3*^M^ mice (**Figure 5H**). Also, flow analysis of spleen and bone marrow showed a similar leukemia immunophenotype as peripheral blood (**Figure 5H**). These results indicate that *JAK3*^*M*^*RUNX1*^*M*^ induced the ETP-ALL phenotype *in vivo*. There was also a trend for higher Mac1+ cells with lower level of CD3+ cells in the peripheral blood of *RUNX1*^M^ mice but they never developed leukemia within this timeframe (**Figure S13**).

## Discussion

RUNX1 plays significant roles in definitive hematopoiesis by regulating the differentiation of myeloid, megakaryocyte, and lymphoid lineages. In this study, we comprehensively investigated *RUNX1* variants in germline ALL samples from patients and identified highly deleterious germline *RUNX1* variants in T-ALL cases, most of which were frameshift or nonsense variations. By multilayer functional experiments and comprehensive epigenomic and genomic profiling analyses, we systematically characterized *RUNX1* variant functions and identified *JAK3* mutations as predominant co-operating somatic lesions in T-ALL. Furthermore, *RUNX1* variant, in conjunction with mutant *JAK3*, directly gave rise to ETP-ALL *in vivo*. These findings advance our understanding of the role of *RUNX1* in the predisposition to childhood ALL.

As a crucial transcription factor that regulates the hematopoietic differentiation of multiple lineages, *RUNX1* is one of the most frequent target genes of chromosomal translocation, mutation, and copy number alteration in different hematopoietic diseases and leukemia. *RUNX1* germline variants are associated with familial platelet disorder with associated myeloid malignancy (FPDMM, OMIM #601399), also known as FPD or FPD/AML (13, 14, 16, 24). Although most patients with FPD progress to myeloid malignancies, ALL has been reported in a minority of cases (14, 15). In MDS and AML with germline *RUNX1* variants, somatic *RUNX1* mutations are the most frequently observed genomic alteration, suggesting they are one of the cooperating events for leukemia progression (18, 25). Other studies identified somatic mutations in *CDC25C, GATA2, BCOR, PHF6, JAK2, DNMT3A, TET, ASXL1* albeit with lower frequencies (18, 25, 26). By contrast, we identified *JAK3* mutations as the predominant co-occurring event with *RUNX1* germline variants in T-ALL, which consistently drove an ETP phenotype in patients and in mouse models. Therefore, we postulate that while germline *RUNX1* variants disrupt normal hematopoiesis and generally increase the risk of leukemia, the lineage specification of these hematological malignancies is mostly dictated by secondary mutations acquired later in life.

Activating *JAK3* mutations have been reported in T-ALL (23). *In vivo* studies using a murine bone marrow transplantation model showed that *JAK3* mutations in the pseudo-kinase domain caused T-cell lymphoproliferative disease that progressed to T-ALL, mainly by increasing the CD8+ cell population (27, 28). This is in line with our observation that *JAK3*^M^ mice exhibited a significant accumulation of CD8+ cells in thymus, peripheral blood, spleen, and bone marrow. However, *JAK3*^M^*RUNX1*^M^ mouse developed lymphoid leukemia with a completely distinctive phenotype which recapitulated human ETP T-ALL features similar to previous reported ETP-ALL mouse models (*e*.*g*., circulating leukemic cells expressed the myeloid cell marker Mac1, but not the lymphoid markers CD8/CD3)(29, 30). Also, the CD4-CD8- (DN), especially DN1 population was particularly enriched in thymocytes from *JAK3*^M^*RUNX1*^M^ mice, as compared with *JAK3*^M^ mice. Alongside genomic findings in T-ALL patients, these *in vivo* experiments indicate that *RUNX1* dominant-negative variants plus *JAK3*-activating mutations most likely result in the ETP T-ALL. A recent study by Brown et al. comprehensively described the genomic landscape of *RUNX1*-related FPD and myeloid malignancy from 130 families (18). In this cohort, missense and truncating germline *RUNX1* variants were equally represented. While truncating variants occurred in both the RUNT domain and the activation domain, missense variants were largely restricted to the DNA-binding RUNT domain. This pattern is significantly different from that in the lymphoid malignancies as described herein. In T-ALL, deleterious *RUNX1* variants were predominantly nonsense or frameshift and the only missense variant resided in the activation domain. Unfortunately, we do not have family history for children with T-ALL carrying *RUNX1* germline variants, and therefore cannot ascertain the exact penetrance on leukemia or FPD. However, given the profound effects on RUNX1 activity and a range of phenotypes *in vitro* and *in vivo*, these variants are likely to be pathogenic. In fact, the Y287* variant seen in our T-ALL cohort has been previously linked to FPD and functional characterization indicated that this variant causes defective megakaryocyte differentiation in the iPSC model (31). In B-ALL, almost all variants were missense and localized outside of RUNT domain, likely with little effects on RUNX1 activity. Although these variants showed little effects on RUNX1 transcriptional activity level, some of them were predicted to be damaging variants by polyphen2 and SIFT (Table 1). More comprehensive functional assays might be needed to definitively determine the effects of these variants.

Genome-wide patterns of RUNX1 binding have been investigated extensively using ChIP-seq assays (21, 32, 33), but there is a paucity of studies directly examining target genes of variant RUNX1. When this was attempted in the past, variant RUNX1 was either ectopically expressed in iPSC or cord blood CD34+ cells, raising the possibility of false positives due to artificially high levels of RUNX1(34). This is also hindered by the lack of antibodies that specifically recognize wildtype but not variant RUNX1. To overcome these issues, we engineered Jurkat cells with heterozygous knock-in of *RUNX1* variants (p.R232fs/WT, p.Y287*/WT, and p.G365R/WT) using the CHASE-KI method (35). In this model, we also introduced the HA and TY1 epitope tags at the 3’ end of the coding exon on the variant and WT *RUNX1* allele, respectively. Our model recapitulated *RUNX1* variant status in patients with T-ALL and enabled us to profile variant or wildtype RUNX1 binding using different antibodies. Moreover, our ChIP-seq result indicated that the C-terminal truncating variants p.R232fs and p.Y287* variants actually exhibited a distinct binding pattern than the full-length p.G365R variant and wildtype RUNX1.

In summary, we comprehensively described *RUNX1* germline variants in childhood ALL. Using multiple functional assays, we identified highly deleterious germline variants in T-ALL and their biochemical and cellular effects. In addition, we characterized somatic genomic alterations associated with *RUNX1* germline variation in T-ALL, illustrating the interplay between acquired and inherited genetic variants in the context of leukemia pathogenesis.

## Methods

### Patients

A total of 6,190 ALL cases were included for RUNX1 targeted sequencing: 4,132 children with newly diagnosed B-ALL enrolled on the Children’s Oncology Group (COG) AALL0232 (n = 2,224), P9904/5/6 (n = 1,634), and AALL0331 (n = 274) protocols; 704 children with newly diagnosed B-ALL enrolled on the St.Jude Total XIII and XV protocols; 1,231 children with newly diagnosed T-ALL enrolled on the COG AALL0434 protocols (1,231); and 123 children with newly diagnosed T-ALL enrolled on the St.Jude Total XIII and XV protocols (Figure 1A) (36-39). This study was approved by institutional review boards at SJCRH and COG affiliated institutions and informed consent was obtained from parents, guardians, or patients, and assent from patients, as appropriate. Family histories were not available for patients on the COG studies, and thus ALL cases were considered to be sporadic. Germline DNA was extracted from peripheral blood or bone marrow from children with ALL during remission.

For targeted *RUNX1* sequencing in the sporadic ALL cohort, Illumina dual-indexed libraries were created from the germline DNA of 6,190 children with ALL, and pooled in sets of 96 before hybridization with customized Roche NimbleGene SeqCap EZ probes (Roche, Roche NimbleGen, Madison, WI, USA) to capture the *RUNX1* genomic region. Quantitative PCR was used to define the appropriate capture product titer necessary to efficiently populate an Illumina HiSeq 2000 flowcell for paired-end 2×100 bp sequencing. Coverage of at least 20-fold depth was achieved across the targeted RUNX1 locus for 99.2% of samples. Sequence reads in FASTQ format were mapped and aligned using the Burrows-Wheeler Aligner (BWA)(40, 41), and genetic variants were called using the GATK pipeline (version 3.1)(41), as previously described, and annotated using the ANNOVAR program(42) with the annotation databases including RefSeq(43), Polyphen2(44, 45) and SIFT(46). Non-coding, and synonymous coding variants were excluded from further consideration for this study.

### Genomic Analysis of Patient Samples

Whole-genome sequencing and RNA sequencing were performed for T-ALL cases with germline RUNX1 variants, whenever available samples were identified. Whole genome seq was done for matched germline and leukemia samples, whereas RNA-seq was done only for leukemia samples. Briefly, DNA was purified using the QIAamp DNA Blood Mini Kit (Qiagen, 51104), and RNA was purified using the RiboPure RNA Purification Kit (Thermo Fisher Scientific, AM1928). DNA (250-1000 ng) and RNA (500-1000 ng) was sent to St. Jude Hartwell center for sequencing. Details for functional experiments, leukemia modeling in mouse, genomic analyses, and other experiments can be found in supplemental file.

More detailed methods can be found in supplemental files.

## Data Availability

All the data in this manuscript are available.

## Notes

### Competing Interest Statement

The authors have declared no competing interest.

### Clinical Trial

St Jude Total studies: NCT00137111
AALL0434: NCT00408005
AALL0232: NCT00075725
AALL0331: NCT00103285
P9904: NCT00005585
P9905: NCT00005596
P9906: NCT00005603

### Funding Statement

No external funding was received.

### Author Declarations

The IRB was approved by the Institutional Review Board of St Jude Children's Research Hospital

