## Supplemental Figures, methods, tables for "Germline *RUNX1* Variation and Predisposition to Childhood Acute Lymphoblastic Leukemia"

### Supplemental information

### Supplemental Figures

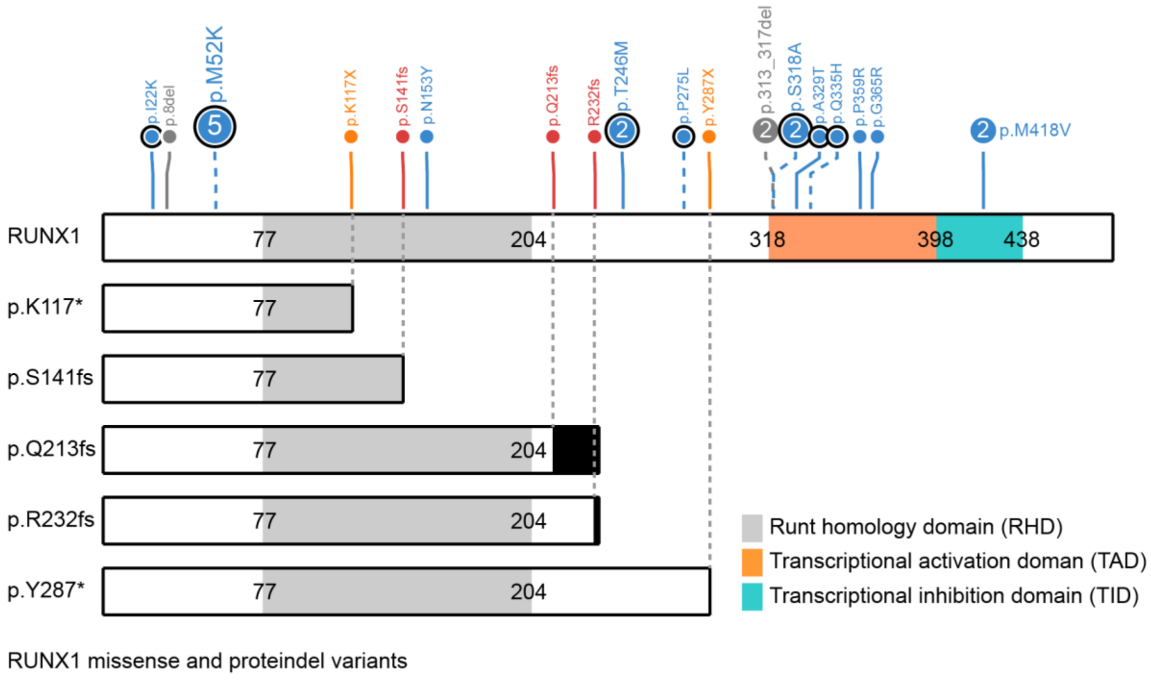

**Figure S1. Schematic diagram of frameshift and nonsense variants in *RUNX1* observed in childhood T-ALL.** p.K117* and p.S141fs variants truncate both the DNA binding domain (RHD) and the transcriptional activation domain (TAD). The p.Q213fs, p.R232fs, and p.Y287* variants truncate the transcriptional activation domain.

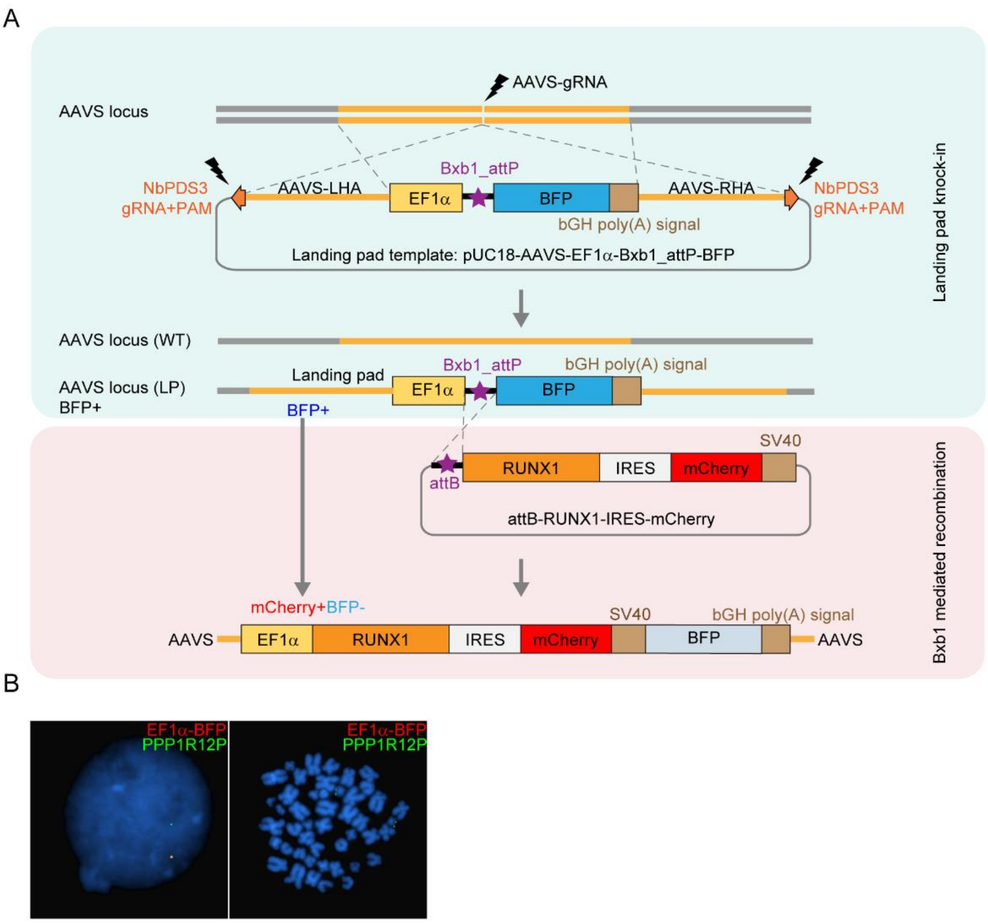

**Figure S2. The design and validation of landing-pad insertion in Jurkat.** (A) Schematic representation of landing pad and Bxb1 mediated attP recombination system. Upper panel: CRISPR-Cas9 mediated homology recombination was used to knock in the landing pad into the *AAVS1* locus, the Bxb1_attP recombination site is juxaposed with BFP coding sequence under control of the EF1α promoter. Landing pad inserted cells are BFP positive. Lower panel: RUNX1-IRES-mCherry cassestte was inserted into the Bxb1_attP site between the EF1α promoter and BFP upon the co-transfection of Bxb1 expression plasmid. The succesfully recombined cells are mCherry$^{+}$BFP$^{-}$. (B) FISH confirmed that a single copy of the landing pad was inserted at the *AAVS1* locus.

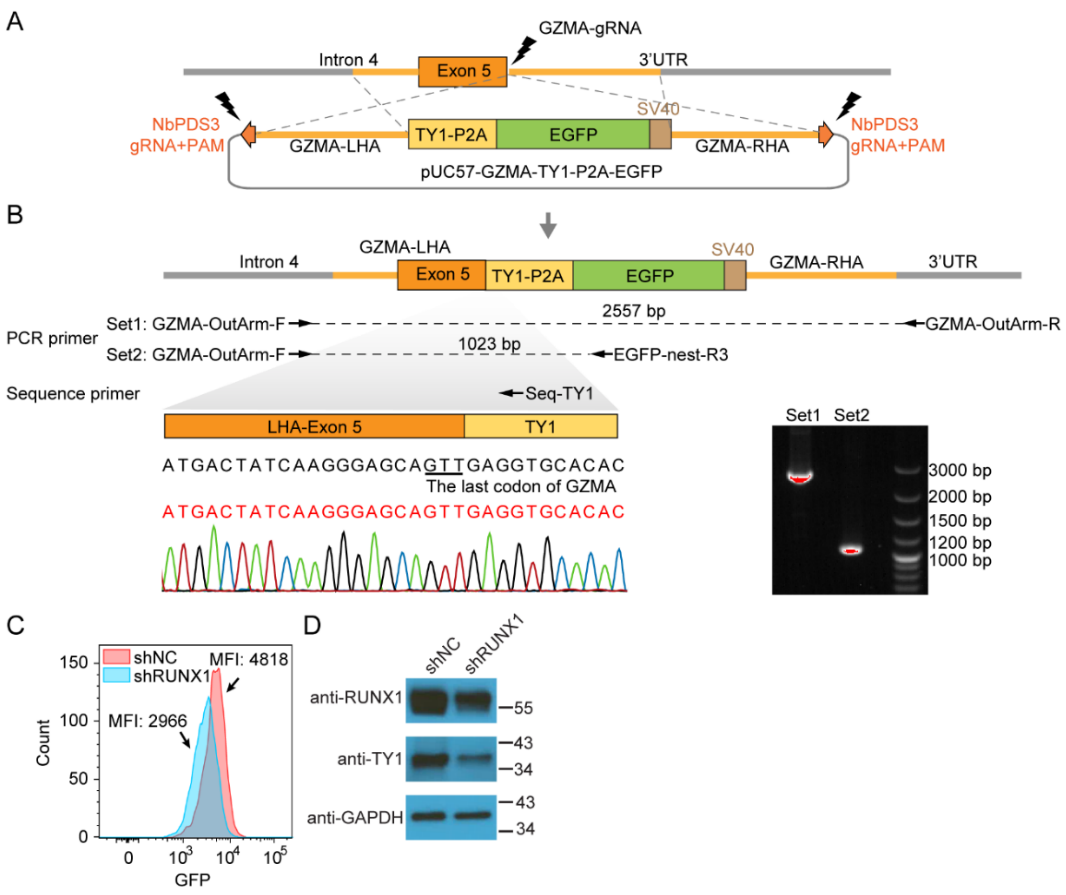

**Figure S3. The design and validation of the EGFP knock-in after *GZMA* coding region in Jurkat cell.** (A) The knock-in of EGFP after *GZMA* coding region was performed using CRISPR-Cas9 mediated homology recombination. TY1-P2A-EGFP was inserted at the endogenous *GZMA* locus before the stop codon. (B) PCR and sanger sequencing result of EGFP knock-in single clone 18 (sc18). Both the sanger sequencing (lower left panel) and PCR (lower right panel) showed TY1-P2A-EGFP-SV40 cassette was inserted homozygously. (C) Flow cytometry showed GFP signal (which reflects GZMA expression level) in the Jurkat cells. When RUNX1 was knocked down using shRNA, a significant decrease was observed in GFP intensity, confirming the effects of RUNX1 on GZMA transcription. (D) Similarly, immonoblotting assay showed that GZMA expression level was decreased after the shRNA knock-down of RUNX1 (detected by TY1 antibody).

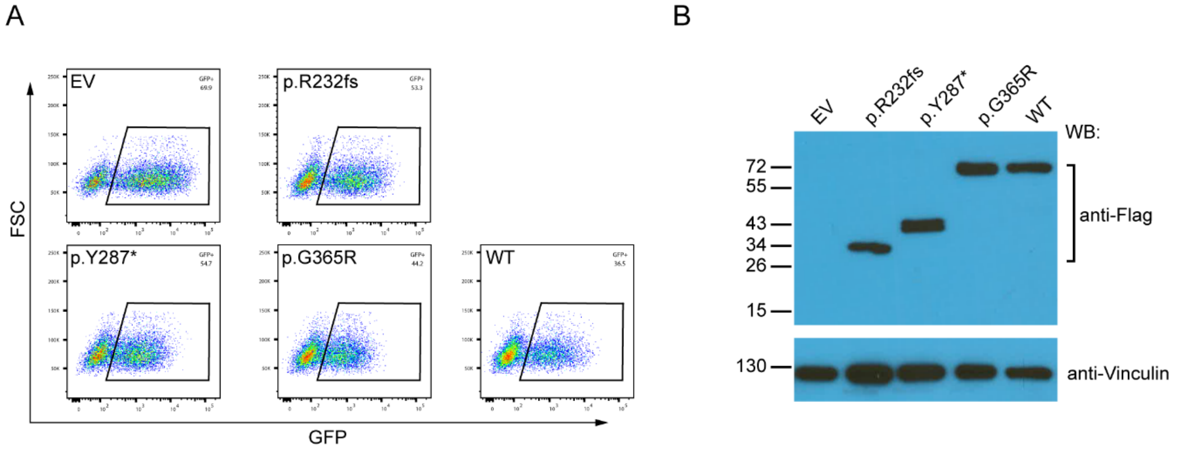

**Figure S4. Ectopic expression of RUNX1 variants in human cord blood CD34+ cells.**

(A) Flow cytometry analysis of human cord blood CD34+ cells transduced with *RUNX1* variants, wildtype *RUNX1* (WT), or empty vector (EV). (B) Western blot confirmed the expression of RUNX1 variants in human cord blood CD34+ cells, vinculin is used as internal control.

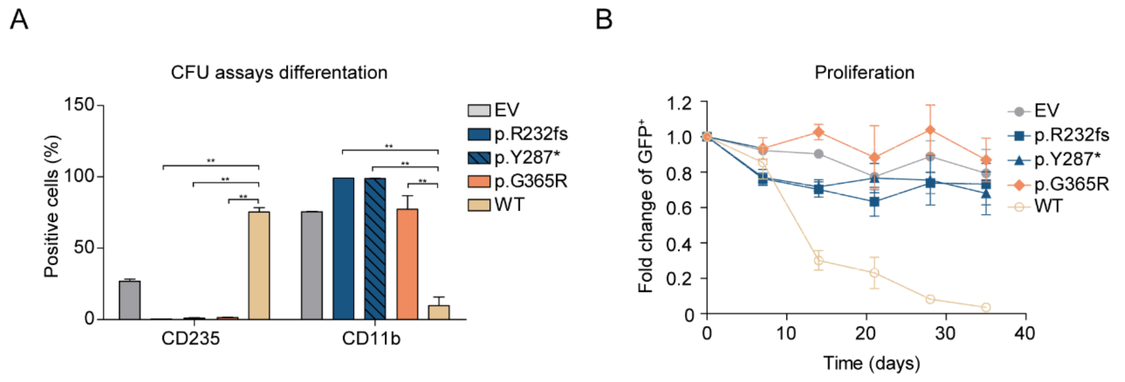

**Figure S5. Immunophenotype of CD34+ cells expressing *RUNX1* variants and proliferation.** (A) Population of erythroid cells (CD235+) and myeloid cells (CD11b+) on CFU assay plates. (B) Changes in the GFP$^+$ population in unsorted samples during long-term culture of human cord blood CD34+ cells transduced with *RUNX1* variants.

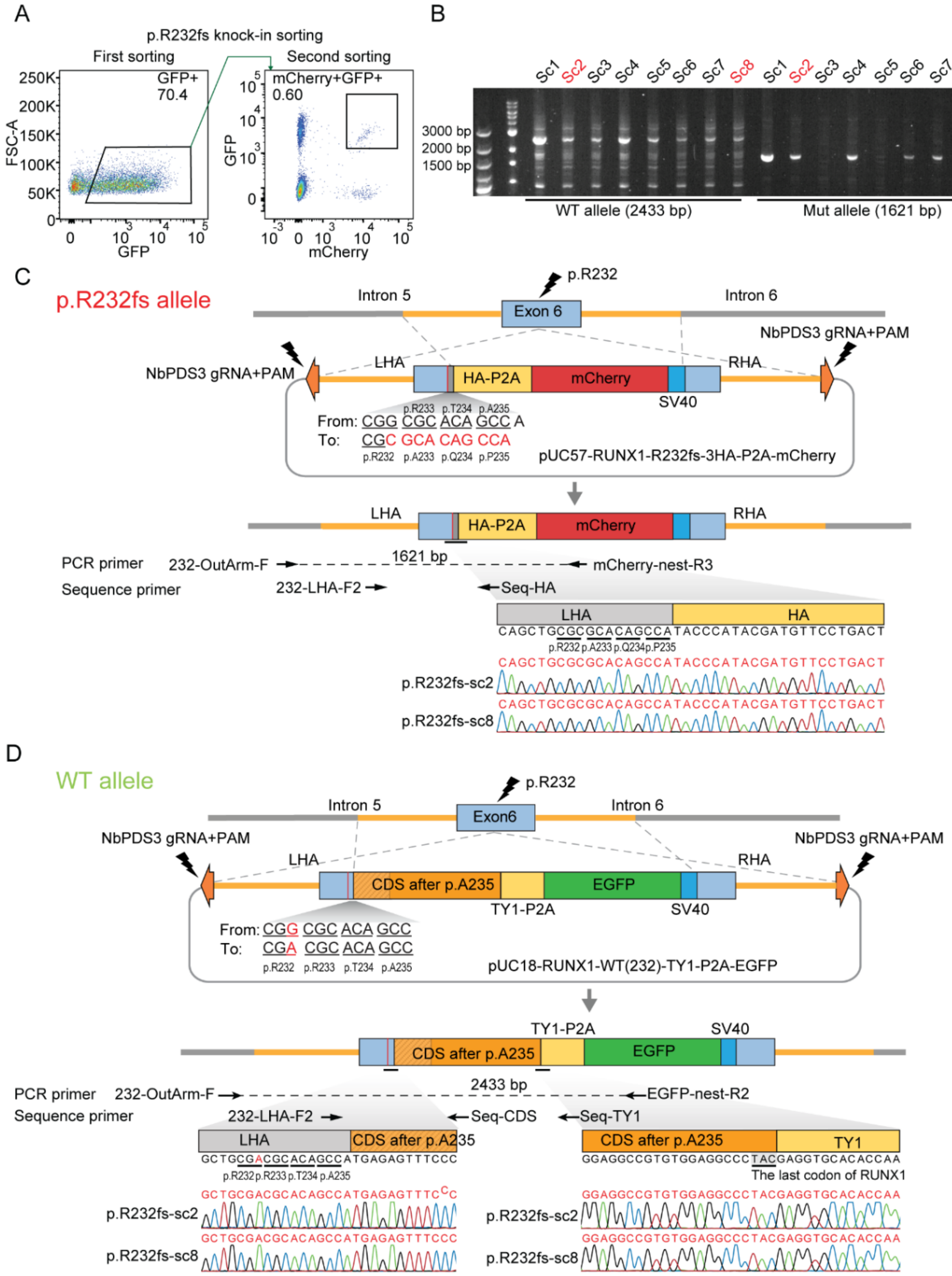

**Figure S6. Design and validation p.R232fs heterozygous knock-in by CRISPR mediated homology recombination.** (A) The mCherry+/GFP+ cells are enriched after two rounds of sorting. (B) DNA gel shows the PCR products of primer sets 232-OutArm - F/mCherry-nest-R3 (1621 bp, mutation allele) and 232-OutArm-F/EGFP-nest-R2 (2433 bp,

WT allele) for eight single clones (we choose sc2 and sc8 in the following experiments). (C-D) *RUNX1* gene locus and the design of p.R232fs knock-in donor plasmid (C) and WT donor plasmid (D) are shown the upper panel. Primer design and Sanger sequencing results for both variant (C) and WT (D) were shown in the lower panel. For p.R232fs allele (C), The HA-P2A-mCherry-SV40 cassette were added after p.R232-p.A233-p.Q234-p.P235, which is identical to the coding change resulted from p.R232fs in patients. For WT allele (D), the LHA and RHA are similar with that of p.R232fs with minor modification as shown in the middle panel.

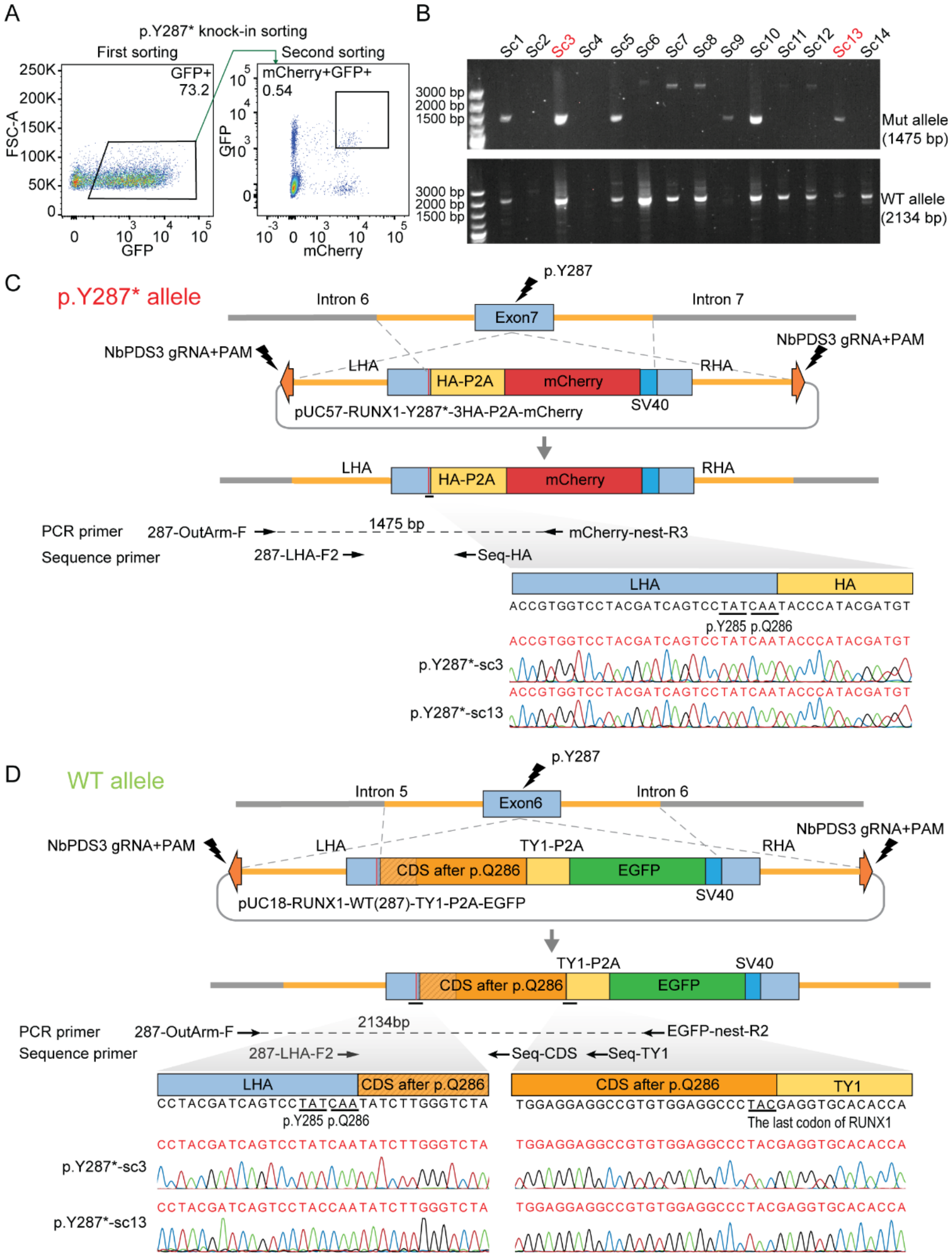

**Figure S7. Design and validation p.Y287* heterozygous knock-in by CRISPR mediated homology recombination.** (A) The mCherry+/GFP+ cells are enriched after two rounds of sorting. (B) DNA gel shows the PCR products of primer sets 287-OutArm -F/mCherry-nest-R3 (1475 bp, mutation allele) and 287-OutArm-F/EGFP-nest-R2 (2134 bp, WT allele) for eight single clones (we choose sc3 and sc13 in the following experiments). (C-D) *RUNX1* gene locus and the design of p.Y287* knock-in donor plasmid (C) and WT donor plasmid (D) are shown the upper panel. Primer design and Sanger sequencing results for both variant (C) and WT (D) were shown in the lower panel. For p.Y287* allele (C), The HA-P2A-mCherry-SV40 cassette were added after p.Q286, which is identical to the coding change resulted from p.Y287* in patients. For WT allele (D), the LHA and RHA are similar with that of p.Y287* with minor modification as shown in the middle panel.

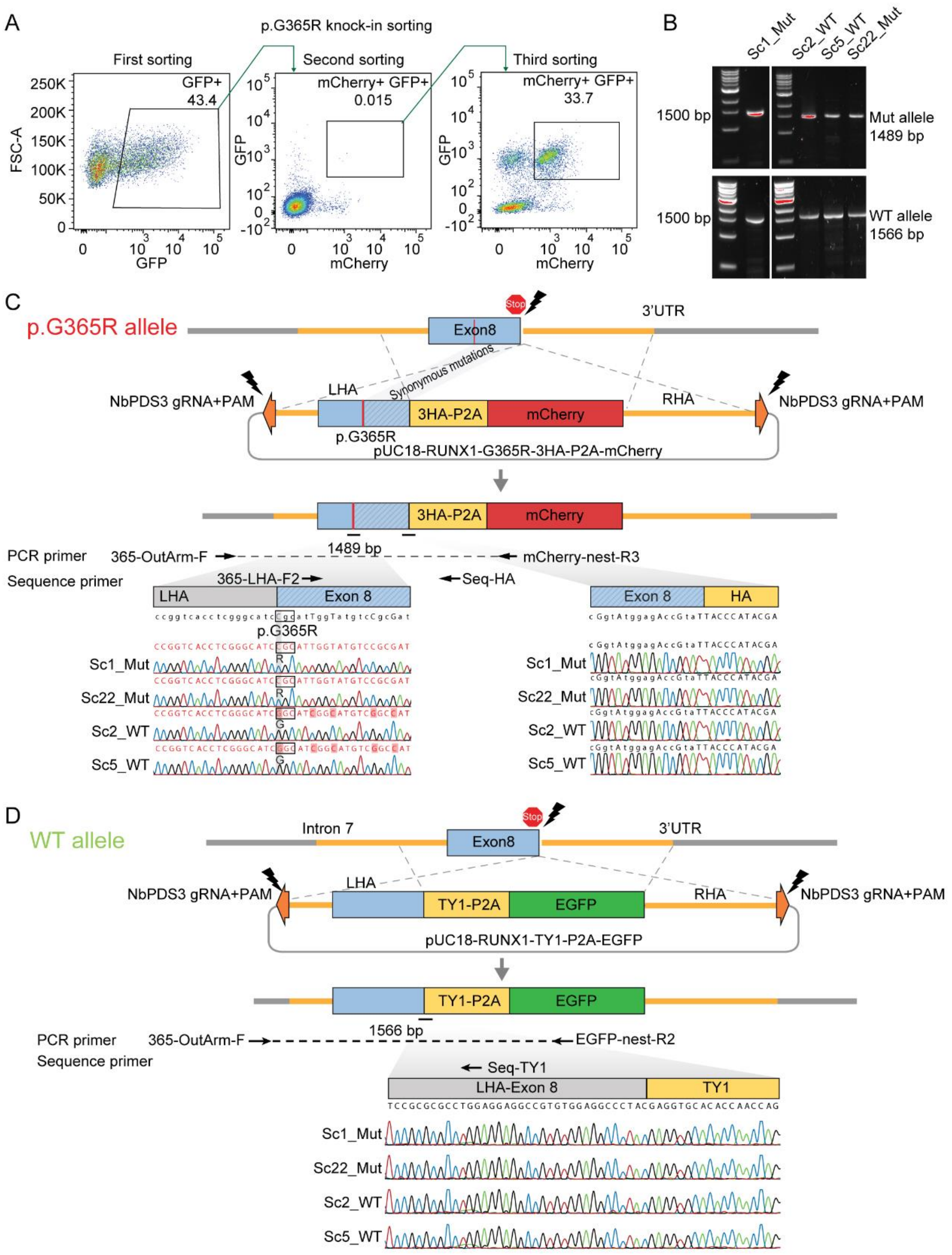

**Figure S8. Design and validation p.G365R heterozygous knock-in by CRISPR mediated homology recombination.** (A) A three-step sorting strategy to enrich cells with successful editing, *i.e.*, mCherry+/GFP+ cells. (B) DNA gel shows the PCR products of primer sets 365-OutArm -F/mCherry-nest-R3 (1489 bp, mutation allele) and 365-OutArm-F/EGFP-nest-R2 (1566 bp, WT allele) for four single clones (p.G365R: sc1 and sc22, WT: sc2 and sc5). (C-D) *RUNX1* gene locus and the design of p.G365R knock-in donor plasmid (C) and WT donor plasmid (D) are shown the upper panel. Primer design and Sanger sequencing results for both variant (C) and WT (D) were shown in the lower panel. For p.G365R allele (C), The p.G365R mutation were generated on LHA, which is identical to the coding change resulted from p.G365R in patients. For WT allele (D), the LHA and RHA are similar with that of p.G365R with minor modification as shown in the middle panel. Sanger sequencing results of two variant clones (sc1 and sc22,) and two WT clones (sc2 and sc5, both alleles expressed WT RUNX1 with either HA or TY1 tag).

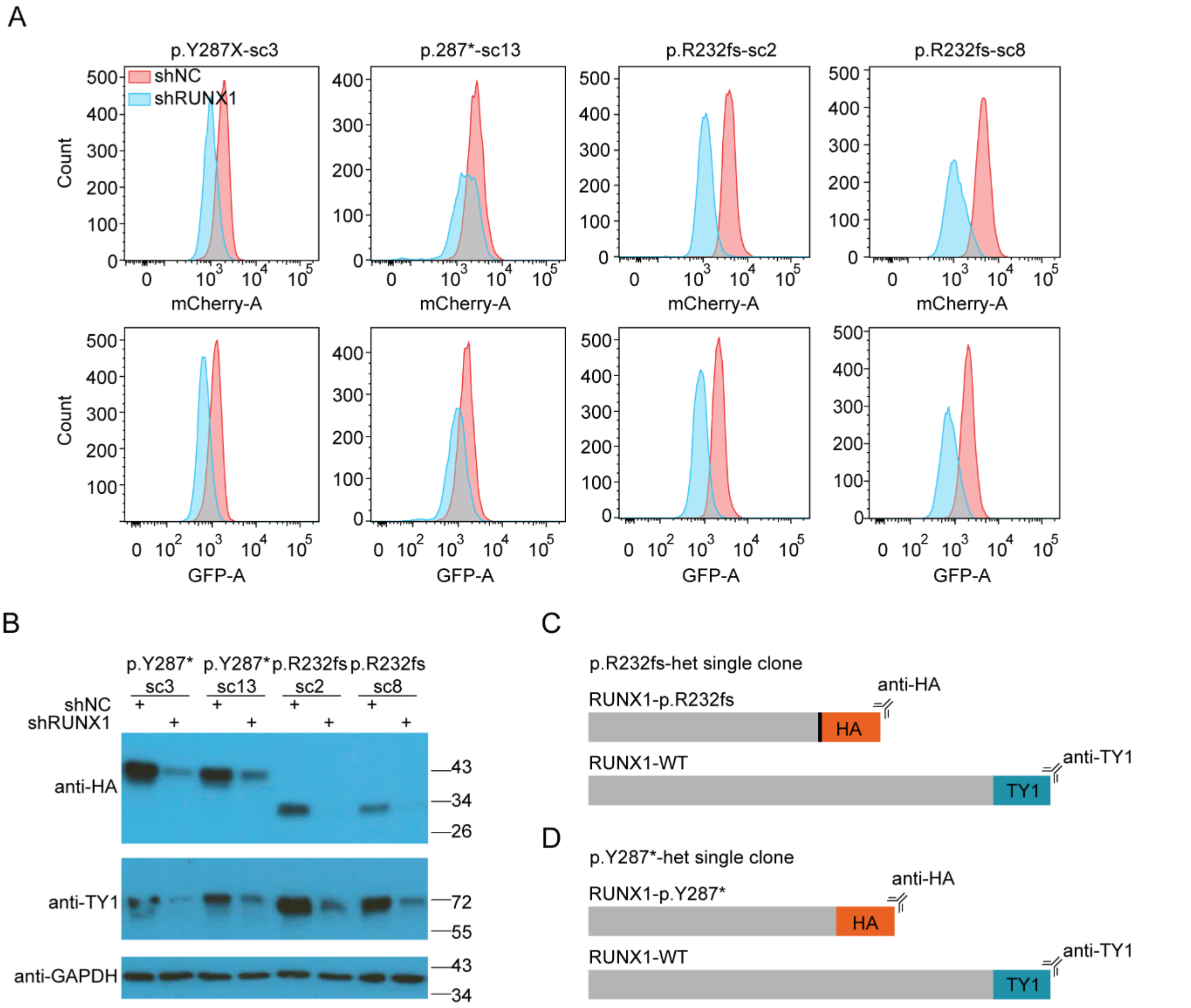

**Figure S9. Validation of heterozygous knock-in of the RUNX1 p.R232fs and p.Y287* variants.** (A) Flow cytometry shows that both mCherry and GFP signals were reduced after RUNX1 knock-down. (B) Immunoblot shows that the expression of both HA- and TY1-tagged RUNX1 were reduced after RUNX1 knockdown. RUNX1 protein levels were detected by anti-HA and anti-TY1 antibodies separately. GAPDH was used as internal control. (C and D) In the p.R232fs-het (C) and p.Y287*-het (D) clones, p.R232fs or p.Y287* alleles were tagged with HA, and WT allele was tagged with TY1.

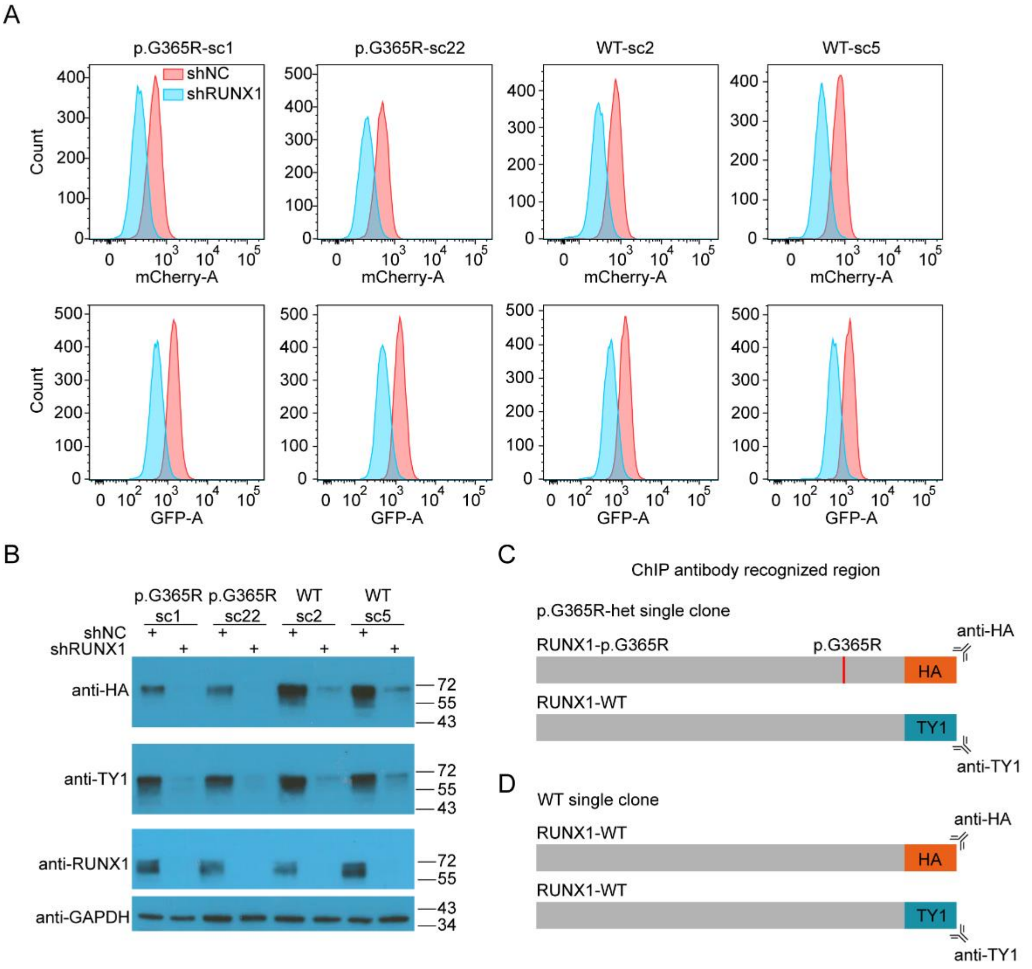

**Figure S10. Validation of heterozygous knock-in of the RUNX1 p.G365R variant.** (A) Flow cytometry shows that both mCherry and GFP signals decreased after RUNX1 knock-down. (B) Similarly, immunoblot confirmed that the expression of both HA- and TY1-tagged RUNX1 were reduced after RUNX1 knockdown. RUNX1 protein levels were detected by anti-HA, anti-TY1, and anti-RUNX1 antibodies separately. GAPDH was used as internal control. (C-D) Schematic diagram of the RUNX1 region recognized by the antibodies used for ChIP. In the p.G365R-heterozygous (p.G365R-het) clones (C), the p.G365R allele is tagged by HA, and WT is tagged by TY1. In the WT clones (D), both RUNX1 alleles are WT and tagged by HA or TY1 separately.

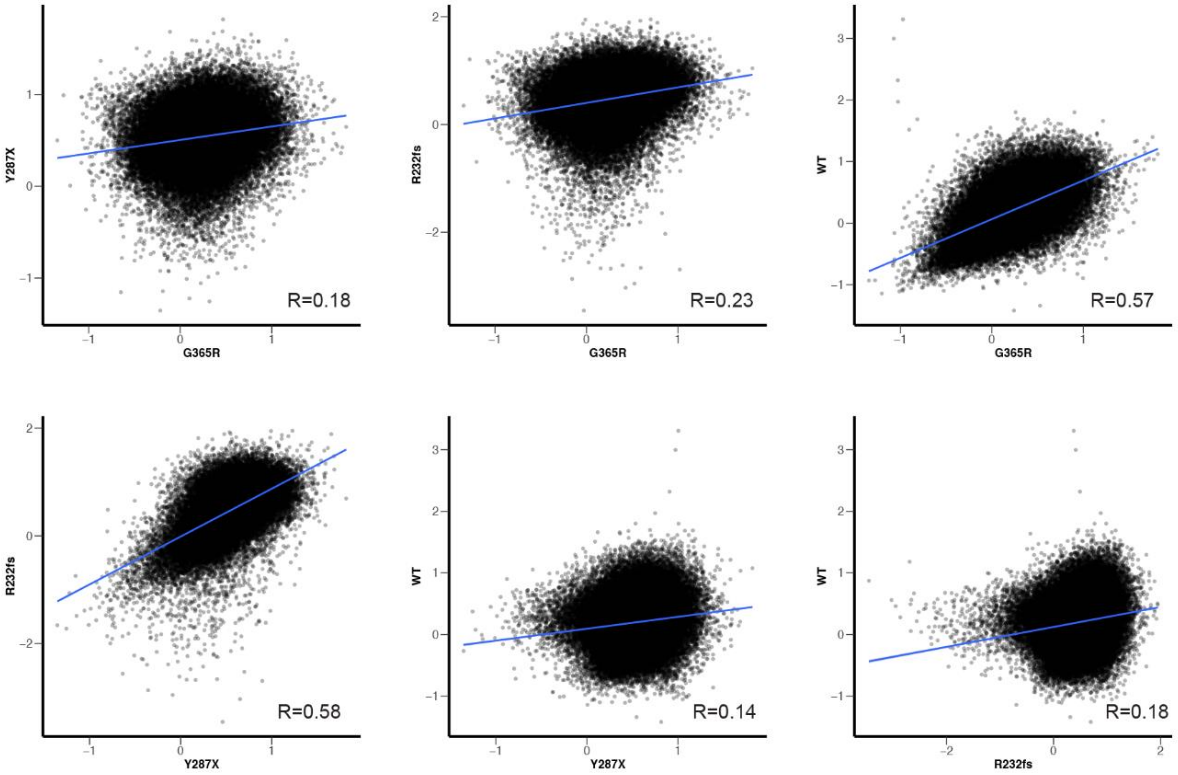

**Figure S11. Scatter plot of WT and variant RUNX1 ChIP-seq signals**. Each X-axis and Y-axis represent the log2 ratio of ChIP-seq signals between HA (WT and variant RUNX1) and TY1 (WT).

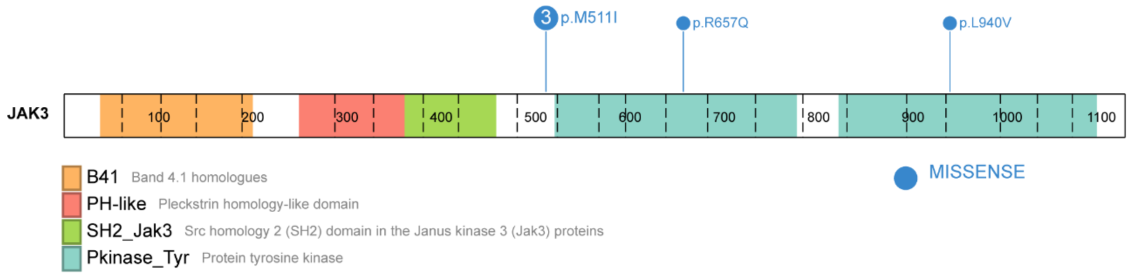

**Figure S12. Somatic *JAK3* mutations identified in seven T-ALL cases containing germline *RUNX1* variants.**

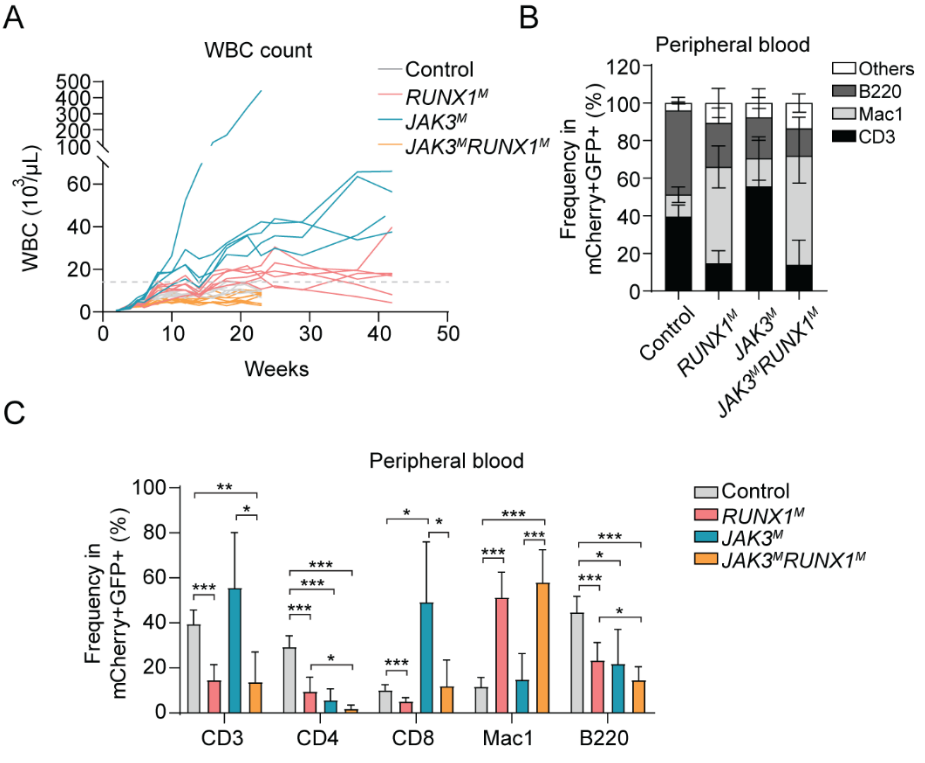

**Figure S13. Leukocyte count and flow cytometry analysis of peripheral blood cells weekly or after 4 months of transplantation.** (A) Peripheral leukocyte count of *RUNX1*$^{M}$, *JAK3*$^{M}$, *JAK3*$^{M}$*RUNX1*$^{M}$, and control mice. Hematopoietic stem and progenitor cells were first lentivirally transduced with *RUNX1*$^{M}$, *JAK3*$^{M}$, *RUNX1*$^{M}$*/JAK3*$^{M}$, or empty vector, which were then injected into recipient animals. Complete blood count (CBC) test of peripheral blood was monitored every other week. (B-C) There was a significant increase in the CD8+ population in *JAK3*$^{M}$ mice, compared with control mice (EV). *RUNX1*$^{M}$ and *JAK3*$^{M}$*RUNX1*$^{M}$ mice showed increases in the Mac1+ population and decreases the CD3+ population as compared with control or *JAK3*$^{M}$ mice after 4 months of transplantation. Control group (EV): n = 7; *RUNX1*$^{M}$ group: n = 8; JAK3$^{M}$ group: n = 5; *JAK3*$^{M}$*RUNX1*$^{M}$ group: n = 6.

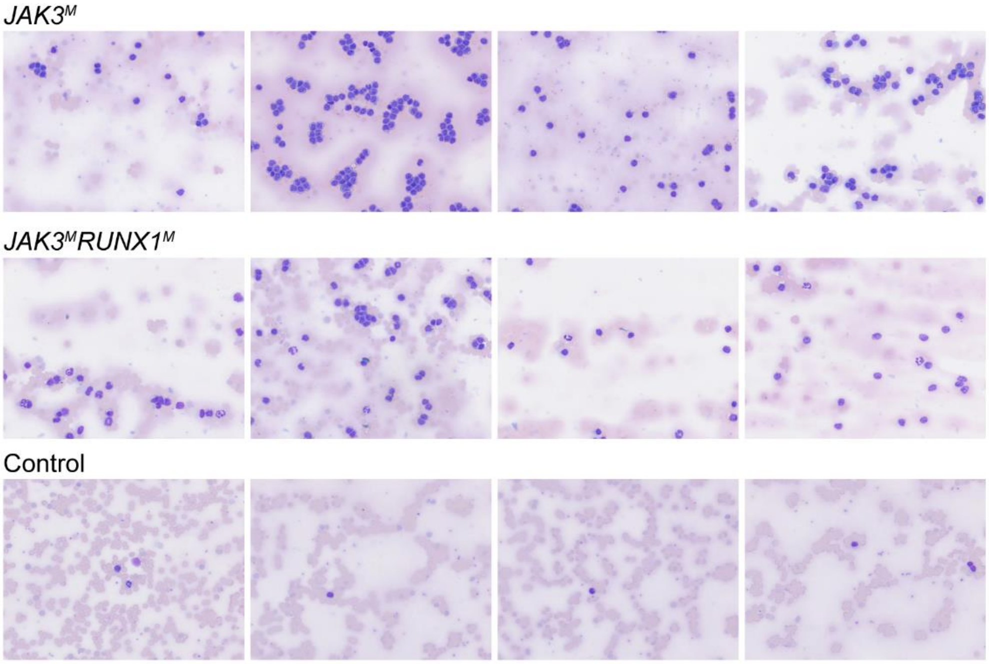

**Figure S14. Examples of blood smear of four *JAK3$^{M}$*and four *JAK3$^{M}$RUNX1$^{M}$* mice at the time of sacrifice, and four control B6 mice after 4 months of transplant.**

### Supplemental Methods

#### Cells and Cell culture

Jurkat cells were purchased from the American Type Culture Collection (ATCC) and cultured in RPMI-1640 containing 10% fetal bovine serum (FBS). Lenti-X 293 cells were purchased from Clontech and cultured in Dulbecco’s Modified Eagle’s Medium (DMEM) with high glucose (4.5 g/L), 4 mM L-glutamine, sodium bicarbonate, 10% FBS, 100 units/mL penicillin G sodium, 100 µg/mL streptomycin sulfate, and 1 mM sodium pyruvate. HEK-293T cells were purchased from ATCC and cultured in DMEM containing 10% FBS. Hela cells were purchased from ATCC and cultured in Eagle's Minimum Essential Medium (EMEM) containing 10% FBS. Human cord blood CD34+ cells were purchased from STEMCELL Technologies and cultured in serum-free expansion medium (SFEMII; STEMCELL, 09605) containing human CD34+ cell–expansion supplement (STEMCELL, 02691).

#### Luciferase Reporter Gene Assay

Human *RUNX1b* cDNA was cloned into the BamHI and XhoI-digested pcDNA3.1 backbone by using NEBuilder® HiFi DNA Assembly Master Mix [New England Biolabs (NEB), E2621]. *RUNX1* variants were introduced using QuikChange II Site-Directed Mutagenesis Kit (Agilent, 200523). Human *CBFβ* cDNA was cloned into the KpnI and XhoI-digested pcDNA3.1 backbone by using NEBuilder HiFi DNA Assembly Master Mix. We cloned the *SPI1* promoter and enhancer region into the pGL3 luciferase-reporter plasmid. Then we transiently transfected Hela cells with *RUNX1* expression plasmid, either WT or a variant, and CBFβ-expression plasmid, SPI1 promoter-driven luciferase reporter plasmid, and SV40 promoter-driven luciferase reporter plasmid as a control. Luciferase activity was measured 24 hours after transfection. Detailed plasmid sequence information can be found in the supplemental file.

**GZMA Reporter Gene Assay**

*GZMA* is known RUNX1 target gene (1) and is therefore its transcription can reflect RUNX1 variant function. We use modified landing-pad strategy (2) to introduce a single copy of each *RUNX1* variant in each cell. The landing pad construct, i.e., Bxb1 intergrase-mediated attP recombination site followed by BFP, was inserted into the *AAVS* safe harbor locus, and cells were then sorted for $BFP^+$ to identify the population with successful insertion. When co-transfected with Bxb1 expression plasmid and attB-RUNX1-IRES-mCherry plasmid (WT or variants), the recombination will give rise to RUNX1 expression driven by the EF1α promoter. The successfully recombined cells will be identified by flow cytometry as $mCherry^+BFP^-$. In parallel, we inserted EGFP coding sequence at the 3' end of *GZMA* resulting in a fusion protein. GFP signal was normalized to the background GFP signal to indicate the activity of each RUNX1 variant (e.g., loss of function, dominant negative, benign), and cells without *RUNX1* insertion at the *AAVS* locus were included as a negative control.

CRISPR/Cas9 with homology arm and sorting enrichment knock-in protocol (CHASE-KI) (3). For landing pad knock-in, we introduced a cleavage in *AAVS* locus by using guide RNA (gRNA) AAVS1 T2 CRIPR in pX330 (Addgene, #72833)(4) and simultaneously transfected homology-directed repair (HDR) template plasmid into Jurkat cells. The HDR template plasmid (pUC18-AAVS-EF1α-Bxb1_attP-BFP) contained the left *AAVS* homology arm (AAVS-LHA), EF1α-

Bxb1_attP-BFP cassette, and the right AAVS homology arm (AAVS-RHA). AAVS-LHA and AAVS-RHA were amplified from pMK232 (CMV-OsTIR1-PURO) (Addgene, #72834)(4). EF1α was amplified from pEF1α-FB-dCas9-puro (Plasmid #100547)(5). Bxb1_attP-BFP was amplified from dAAVS1–TetBxb1BFP plasmid(2). We added NbPDS3 gRNA plus a PAM sequence at the 5' end of both AAVS-LHA and AAVS-RHA during PCR amplification. These fragments were amplified with 18~20 bp overhang sequence and subcloned into BamHI and KpnI-digested pUC18 backbone by using NEBuilder® HiFi DNA Assembly Master. Human *RUNX1b* cDNA with an N-terminal 2×Flag tag was cloned into the SacI and AflII-digested attB-IRES-mCherry backbone(2) by using NEBuilder® HiFi DNA Assembly Master Mix. *RUNX1* variants were introduced using QuikChange II Site-Directed Mutagenesis Kit.

A total of $5 \times 10^6$ Jurkat cells were transiently transfected with 3 µg pUC18-AAVS-EF1α-Bxb1_attP-BFP, 1 µg AAVS1 T2 CRIPR in pX330, and 1 µg NbPDS3-gRNA. The first sorting for successfully transfected ($GFP^+$) cells was performed 24 hours after transfection by BD Biosciences Aria cell sorters. The second sorting for landing pad knock-in ($BFP^+$) cells were performed 14 days after the first sorting. Single clones were selected, and single copy knock-in were verified by PCR and FISH. We choose one single copy landing pad knock-in cells for the following experiments. All primer, gRNA, and HDR template sequences are provided in the supplemental file.

CHASE-KI (3) was used for the knock-in of the EGFP into the C terminal of endogenous *GZMA* allele before the stop codon. For EGFP knock-in, we introduced a cleavage after the *GZMA* STOP codon by using guide RNA (gRNA) and transfected the *GZMA* HDR template plasmid (pUC57-GZMA-TY1-P2A-EGFP) into Jurkat cells. The sequence of *GZMA* HDR template plasmid is shown in the supplemental file. Briefly, the LHA of the *GZMA* HDR template contains exon 5 and part of intron 4 (from –600 bp to 0 bp upstream towards the *GZMA* STOP codon). The RHA of the *GZMA* HDR template contains the 3'UTR region (600 bp downstream from the *GZMA* STOP codon). We introduced a mutation after 14 bp of STOP codon to destroy the PAM sequence of gRNA recognition region, to eliminate the possibility that gRNA recognizes this region and cut it again after the homology recombination. Between the LHA and RHA is TY1-P2A-EGFP. The HDR template, synthesized by GENEWIZ, is surrounded by NbPDS3 gRNA plus a PAM sequence and are cloned into the pUC57 plasmid by GENEWIZ. We add P2A between *GZMA* and *EGFP*. As a self-cleaving peptide, P2A ensures that EGFP were co-translated with the GZMA but separate from GZMA by the posttranslational cleaving. This design not only ensure the GFP level equals the GZMA expression level, but also eliminate any possibility that GFP influence the GZMA function as fusion protein.

A total of $5 \times 10^6$ Jurkat landing pad knock-in cells were transiently transfected with 3 μg (pUC57-GZMA-TY1-P2A-EGFP), 1 μg px458-GZMA-STOP, and 1 μg NbPDS3-gRNA. The first sorting for successfully transfected ($BFP^+GFP^+$) cells was performed after 24 hours of transfection by BD Biosciences Aria cell sorters. The second sorting for GZMA reporter knock-in ($BFP^+GFP^+$) cells were performed 14 days after the first sorting. Single clones were selected, and knock-in was verified by PCR, Sanger sequencing, and immunoblotting. We choose a homozygous GFP knock-in single clones for the following experiments. All primer, gRNA, and HDR template sequences are provided in the supplemental file.

A total of $3\times10^6$ Jurkat-GZMA-GFP reporter cells were transiently transfected with 4 μg attb-RUNX1-IRES-mCherry and 2 μg of Bxb1 expression plasmid (2) using Amaxa® Cell Line Nucleofector® Kit R, program T-016 (Lonza VVCA-1001). Flow cytometry analysis was performed to quantify GFP intensity in the mCherry$^{+}$BFP$^{-}$ population, as a measurement of RUNX1 activity.

**Fluorescence in Situ Hybridization (FISH) to confirm landing-pad insertion**

Jurkat landing pad knock-in cells were harvested after four hours of colcemid (Thermo Fisher Scientific, 15212012) incubation. Purified puc18-EF1α-BFP plasmid and Ppp1r12c BAC clone (CH17-416P7 / 19q13.42) (BACPAC Genomics Inc) (*AAVS1* locus) were used as the probes for FISH assays. The puc18-EF1α-BFP DNA was labeled with Alexa Fluor™ 594-5-dUTP, C11400) by nick translation and the Ppp1r12c BAC was labeled with Alexa Fluor™ 488-5-dUTP (Fisher Scientific, C11397). Both labeled probes were mixed with sheared human DNA and hybridized to metaphase and interphase nuclei derived from Jurkat landing pad knock-in cells in a solution containing 50% formamide (Millipore, 4610-OP), 10% dextran sulfate (Millipore, S4030), and 2X SSC (Sigma, S6639). The cells were then stained with 4,6-diamidino-2-phenylindole and fluorescence images were taken by Nikon Eclipse 80i. Detailed plasmid sequence information can be found in the supplemental files.

**Fluorescence Microscopy**

Human *RUNX1b* cDNA was fused in-frame with mCherry or EGFP coding sequences and then cloned into BamHI and XhoI-digested pcDNA3.1 backbone by using NEBuilder HiFi DNA Assembly Master Mix. PcDNA3.1-mCherry-RUNX1b was subsequently used to generate

*RUNX1* variant sequence by QuikChange II Site-Directed Mutagenesis Kit. The mCherry-tagged RUNX1b (variants) and EGFP-tagged RUNX1b (WT) were co-transfected into HEK293T cells (Emsdiasum, 72222-01) using lipofectamine 2000 (Thermo Fisher Scientific, 11668030). 24 hours after transfection, cells were fixed with 4% paraformaldehyde (Boster, AR1068). Fluorescence images were taken using a Nikon C2 confocal microscope. Detailed information about plasmid sequence can be found in the supplemental files.

**Co-immunoprecipitation assay for RUNX1-CBFβ interaction**

Human *RUNX1b* cDNA with an N-terminal 2×Flag tag was cloned into the EcoRI-digested cl20c-MSCV-GFP backbone by using NEBuilder® HiFi DNA Assembly Master Mix. *RUNX1* variants were introduced using QuikChange II Site-Directed Mutagenesis Kit. Human HEK-293T cells (seeded at $5 \times 10^6$ cells per 10-cm dish, 24 hours before transfection) were co-transfected with 5 µg pcDNA3.1-CBFβ and 5 µg cl20c-2Flag-RUNX1b-IRES-GFP (WT or variant) by using polyethyleneimine. After 24 hours in culture, cells were collected, washed with 1× PBS, lysed with 1 mL RIPA lysis and extraction buffer (Thermo Fisher Scientific, 89900) containing PMSF (Sigma, 10837091001) and proteinase inhibitor cocktails (Sigma, SRE0055). The cell lysate was then incubated with 20 µL anti–FLAG M2 magnetic beads (Sigma, M8823) for 3 hours, washed with RIPA lysis and extraction buffer 5 times. Finally, the magnetic beads were resuspended with 2× Laemmli sample buffer (Bio-Rad, 161-0737) and boiled at 100 °C for 10 minutes. Protein samples were separated on 4%–15% Mini-PROTEAN TGX™ Precast Protein Gel (Bio-Rad, 4561086) and stained with the following primary antibodies: anti-Flag (CST, 14793S), anti-CBFβ (R&D, AF7349-SP), or anti-RUNX1 (CST, 4334S); followed by staining with one of the following secondary antibodies: VeriBlot for IP Detection Reagent (horseradish peroxidase, HRP) (Abcam, ab131366) or anti–donkey, anti–sheep IgG (H+L), and HRP (Thermo Fisher Scientific, A16041). Detailed plasmid sequence information can be found in the supplemental file.

**Lentivirus Production**

To generate cl20c-MSCV-IRES-mCherry plasmid, mCherry was cloned into BmgBI and NotI-digested cl20c-MSCV-IRES-GFP by using NEBuilder® HiFi DNA Assembly Master Mix. Then human *JAK3* cDNA was amplified from pDONR223-JAK3 (Addgene #23944) and cloned into the EcoRI-digested cl20c-MSCV-IRES-mCherry backbone by using NEBuilder® HiFi DNA Assembly Master Mix. *JAK3-M511I* mutation was introduced using QuikChange II Site-Directed Mutagenesis Kit.

A total of $10^7$ human Lenti-X 293 cells (Takara) were seeded in a 10-mL dish 8 hours before transfection. The following plasmids were mixed with 12-fold (volume/weight) PEI and 100-fold (volume/weight) Opti-MEM (Thermo Fisher Scientific, 31985088): 6 µg pCAGkGP1.1R, 2 µg pCAG-VSVG, 2 µg pCAG4-RTR2, and 15 µg cl20c-MSCV-2Flag-RUNX1(WT or variants)-IRES-GFP [or 17.8 µg cl20c-MSCV-JAK3(M511I)-mCherry, 12.7 µg cl20c-MSCV-IRES-GFP, 12.7 µg cl20c-MSCV-IRES-mCherry]. The mixture was vortexed for 10 seconds and incubated at room temperature for 20 minutes. The DNA-PEI-Opti-MEM mixture was then added dropwise to the plate. Medium was changed 12 hours after transfection. Two days later, the supernatant was harvested, filtered through a 0.45-nm filter, and concentrated 100-fold using ultracentrifugation (Beckman). Primers for plasmid construction can be found in the supplemental file.

#### *In vitro* Differentiation Assay in Human Cord Blood CD34+ Cells

Human cord blood CD34+ cells were thawed and resuspended in SFEMII, cultured in SFEMII containing CD34+ expansion supplement (STEMCELL, 02691) for 5 days. Lentiviral transfection was performed on Day 5. Briefly, CD34+ cells were resuspended in SFEMII at a final concentration of $1\times10^6$ cells/mL and split into 24-well plate (200 μl/ well), then 10 μL preconcentrated lentivirus (100-fold concentrated) was added to each well and maintained in culture at 37 °C. Then 1 mL SFEMII was added after 6 hours of incubation. $GFP^+$ cells were sorted 2 days after transfection. For the CFU assay, 1000 cells were plated in H4034 medium and maintained in culture for 2 weeks. The number of BFU-E, CFU-M, and CFU-GM clones were counted on Day 14. Cells from each plate were then resuspended and stained with anti-CD11b (PE, BD Biosciences, Clone ICRF44) or anti-CD235 (BV650, BD Biosciences, Clone GA-R2). Flow cytometric analysis of the stained cells was performed on the BD LSRFortessa™ cell analyzer (BD Biosciences). Flow data were analyzed on FlowJo_V10. Megakaryocyte-differentiation assays were performed using StemSpan Megakaryocyte Expansion Supplement according to the manufacturer's protocol (STEMCELL, 02696). Briefly, 15,000 $GFP^+$ cells were resuspended in 1 mL SFEMII containing megakaryocyte expansion supplement and then transferred into a 24-well plate. On Day 7, 1 mL SFEMII containing megakaryocyte expansion supplement was added to each well. Flow cytometric analysis was performed on Day 14. Anti–human CD41a (APC, BD Biosciences, Clone HIP8) and anti–human CD42b (PE, BD Biosciences, Clone HIP1) were used for flow cytometry. T-cell progenitor–differentiation assays were performed using StemSpan T-Cell Progenitor Differentiation Kit (STEMCELL, 09900) according to the manufacturer's instructions. Briefly, a 24-well plate was coated with StemSpan Lymphoid Differentiation Coating Material (100-fold diluted in PBS) for 2 hours at room temperature. Then each well was washed with 1× PBS, and 10,000 cells were resuspended in 1 mL SFEMII containing lymphoid progenitor expansion supplement. On Day 4, 1 mL fresh medium was added. On Days 7 and 11, half the medium was replaced with fresh medium. Flow cytometric analysis was performed on Day 14. Anti–human CD5 (APC, BD Biosciences, Clone UCHT2) and anti–human CD7 (PE, BD Biosciences, Clone M-T701) were used for flow cytometry.

To examine the effects of RUNX1 variants on proliferation and apoptosis, RUNX1-expressing CD34+ cells were maintained in culture in Iscove's Modified Dulbecco Medium (IMDM) (STEMCELL, 36150) containing 20% BIT9500 (STEMCELL, 09500) and 10 ng/mL FLT-3 ligand (STEMCELL, 78009.1), TPO (STEMCELL, 2522), SCF (STEMCELL, 78062.1), IL-3 (STEMCELL, 78146), and IL-6 (STEMCELL, 78050.1). The number of cells was counted every week for 5 weeks. Cells were also analyzed by flow cytometry for apoptosis on day 7 and day 16, using Annexin-V and DAPI staining. Unsorted CD34+ cells were maintained in the same culture medium. The GFP+ population was detected by flowcytometry every week for 5 weeks.

#### ***RUNX1* Variant Knock-In in Jurkat cells for ChIP-seq**

CHASE-KI protocol (3) was used to introduce p.R232fs, p.Y287*, and p.G365R variants at the endogenous locus, by homology-based recombination. p.G365R localized on the last exon of *RUNX1*. For p.G365R knock-in, we introduced a cleavage near the *RUNX1* STOP codon by using guide RNA (gRNA) and simultaneously transfected Jurkat cells two different HDR template plasmids. The sequence of HDR template for the p.G365R allele (p.G365R-MUT-LHA-3HA-P2A-mCherry-RHA) and that for the WT allele (p.G365R-WT-LHA-TY1-P2A-EGFP-RHA) are shown in the supplemental file. Briefly, the LHA of the p.G365R HDR template contains exon 8 and part of intron 7 (from –800 bp to 0 bp upstream of the *RUNX1* STOP codon). The codon for p.G365 localized on LHA, and mutated from “GGC” to “CGC”, to generate p.G365R mutation. The RHA of the p.G365R HDR template contains the 3'UTR region (451 bp downstream of the *RUNX1* STOP codon). Between the LHA and RHA is HA-P2A-mCherry. The WT HDR template contains exon 8 and part of intron 7 (starting from –800 bp to 0 bp upstream of *RUNX1* STOP codon). The RHA of the p.G365R HDR template contains the 3'UTR region (800 bp downstream of the *RUNX1* STOP codon). TY1-P2A-EGFP is located between the LHA and RHA.

p.R232fs generates a frameshift on exon 6 and produce a termination codon on after amino acid 235. The sequence of HDR template for the p.R232fs allele (p.R232fs-MUT-LHA-3HA-P2A-mCherry-RHA) and that for the WT allele (p.R232fs-WT-LHA-TY1-P2A-EGFP-RHA) are shown in the supplemental file. Briefly, the LHA of the p.R232fs HDR template contains part of intron 5 and part of exon 6. The sequence of LHA was modified according to p.R232fs to make sure that the homology recombination introduces p.R232fs in the knock-in allele. The RHA of the p.R232fs HDR template contains the rest part of exon 6 and part of intron 6 (797 bp downstream of the stop codon generated by p.R232fs). Between the LHA and RHA is HA-P2A-mCherry-SV40 poly(A). The LHA of the WT HDR template is similar with that of p.R232fs without the aforementioned modification. To eliminate the possibility of gRNA-mediated cleavage once recombination occurs, we destroyed the gRNA-PAM sequence on LHA by introducing a synonymous mutation. The RHA of the WT HDR template is same with that of p.R232fs. We add the rest of *RUNX1* coding sequence after the LHA to generate a full-length WT *RUNX1* CDS. Between the LHA and RHA of WT HDR templates is “RUNX1-CDS after p.A235”-TY1-P2A-EGFP-SV40 poly(A).

p.Y287* localized on exon 7. The sequence of HDR template for the p.Y287* allele (p.Y287*-MUT-LHA-3HA-P2A-mCherry-RHA) and that for the WT allele (p.Y287*-WT-LHA-TY1-P2A-EGFP-RHA) are shown in the supplemental file. Briefly, the LHA of the p.Y287* HDR template contains part of intron 6 and part of exon 7 (from –800 bp to 0 bp upstream of the third base of Q286). To eliminate the possibility of gRNA-mediated cleavage once recombination occurs, we destroyed the gRNA-PAM sequence on LHA by introducing a synonymous mutation. The RHA of the p.Y287* HDR template contains part of exon 7 and part of intron 7 (797 bp downstream of Q286). Between the LHA and RHA of p.Y287* HDR templates is HA-P2A-mCherry-SV40 poly(A). Both the LHA and RHA of the WT HDR template are same with that of p.Y287*. We add the rest of *RUNX1* coding sequence after the LHA to generate a full-length WT RUNX1 *CDS*. Between the LHA and RHA of WT HDR templates is “*RUNX1*-CDS after p.Q286*”-TY1-P2A-EGFP-SV40 poly(A).

All HDR templates, synthesized by GENEWIZ, are surrounded by NbPDS3 gRNA plus a PAM sequence and are cloned into the pUC18 or pUC57 plasmid. Following transfection, both the variant and WT HDR templates were removed from the donor plasmid by NbPDS3 gRNA-generated from a plant sequence with a low likelihood of off-target binding in the human genome- to increase the recombination efficacy. Because mCherry and EGFP were inserted after either variant or WT *RUNX1* coding region. Flow sorting of mCherry$^+$/GFP$^+$ cells was performed to enrich cells with successful knock-in.

To generate heterozygous mutation knock-in cells, a total of $5 \times 10^6$ Jurkat cells were transiently transfected with 3 μg mutation HDR plasmid, 3 μg WT HDR plasmid, 2 μg px458-*RUNX1*-stop-codon/p.R232fs/p.Y287*-gRNA (to generate cleavage on RUNX1 allele), and 1 μg NbPDS3-gRNA (to release HDR template from the donor plasmid). The first sorting for successfully transfected (GFP$^+$) cells was performed after 24 hours of transfection by BD Biosciences Aria cell sorters. The second and third sorting (p.R232fs and p.Y287* have two rounds of sorting, p.G365R have three rounds of sorting) for both allele knock-in (GFP$^+$/mCherry$^+$) cells were performed 14 and 37 days after the first sorting. Single clones were selected, and knock-in of p.G365R, p.R232fs, or p.Y287* was verified by PCR, immunoblotting, and flow cytometry. All primer, gRNA, and plasmid sequences are provided in the supplemental file.

**Chromatin-immunoprecipitation Assays**

Chromatin-immunoprecipitation (ChIP) assays were performed on *RUNX1* variant knock-in Jurkat cells by using ChIP-IT High Sensitivity kit (Activemotif, 53040) according to the manufacturer's protocol. Briefly, a total amount of $2 \times 10^6$ cells were treated with Complete Cell Fixative Solution (provided by ChIP-IT High Sensitivity kit) for 15 minutes and then Stop Solution (provided by ChIP-IT High Sensitivity kit) for 5 minutes, followed by sonication. The following antibodies are used for ChIP assays: anti-HA (Abcam, ab9110), anti-TY1 (Diagenode,

C15200054), and normal rabbit IgG (CST, 2729). Primers for ChIP-qPCR analysis are shown in the supplemental data.

**Immunoprecipitation–Mass Spectrometry**

Human HEK-293T cells (seeded at 5×10$^6$ cells per 10-cm dish for 24 hours before transfection, two dishes per group) were transfected with 10 μg pcDNA3.1-2Flag-RUNX1b (WT or p.G365R mutation) using PEI. After 40 hours of culture, cells were collected, washed with 1× PBS, lysed with 1 mL RIPA lysis and extraction buffer containing PMSF, proteinase inhibitor cocktails, and PhosSTOP (Sigma, PHOSS-RO). Cell lysates were incubated on ice for 1 hour, followed by sonication and centrifugation (14,000 g) for 10 minutes at 4 °C. Supernatants were then incubated with 20 μL prewashed anti–FLAG M2 magnetic beads for 2 hours, washed 5 times with RIPA lysis and extraction buffer. Then 40 μL elution buffer [150 mM NaCl, 50 mM Tris at pH 7.5, 1 mM EDTA, 0.05% NP40, 10% glycerol, 500 μg/mL FLAG peptide (Sigma, F3290)] was added, and supernatant was rotated at room temperature for 30 min. Then 20 μL 4× Laemmli sample buffer was added to the eluent and boiled at 100 °C for 10 minutes. All samples were loaded onto a 4%–15% Mini-PROTEAN TGX™ Precast Protein Gel for 10 minutes and stained using GelCode Blue Stain Reagent (Thermo Fisher Scientific, 24590). All the protein bands were cut from the gel and used for mass spectrometry.

**Murine Bone Marrow Transplantation and Leukemia Modeling**

C57/BL6 mice were purchased from The Jackson Laboratory. Bone marrow cells of female C57/BL6 mice were collected from the femur, tibia, pelvis, and humerus. Lineage-negative c-Kit and Scal-1$^+$ cells were then enriched by flow cytometry and maintained in culture in SFEM

(STEMCELL, 09600) containing 10 ng/mL mSCF (78064), 20 ng/mL IGF2 (78221), 20 ng/mL mTPO (78072.1), 10 ng/mL hFGF, and 5 μg/mL protamine sulfate (Sigma, P3369). Lentiviral transduction was performed using a retronectin- and lentivirus-coated 96-well plate. EGFP$^+$/mCherry$^+$ cells were sorted after 48 hours of transduction, washed with PBS, and injected into the tail vein of sub-lethally irradiated female C57/BL6 mice. Mice received LSK cells transduced with RUNX1-p.R232fs/ or empty vector (GFP$^+$) and JAK3-M511I/ or empty vector (mCherry$^+$). CBC test was performed every two weeks after transplantation. Flow cytometry were performed after 4 months of transplantation or when sacrifice the mice.

**ChIP-sequencing (seq), whole genome-seq, and RNA-seq data analysis**

ChIP-Seq data was mapped to the human genome (GRCh37) by Bowtie2 (ver. 2.2.9) (6) with default parameters. Peak calling was performed by MACS2 (ver. 2.1.1.20160309) (7) with default parameters. Peaks with p-value < 1.0e-5 were reported.

Whole genome-seq was performed for matched germline-leukemia pairs, respectively. For whole genome seq, libraries were constructed using Kapa Hyperprep kit (Roche) according to manufacturer's protocols and sequenced via HiSeq 2000/2500 and NovaSeq 6000 (2 x 151 bp pair-end reads). Whole genome-seq analyses were performed following procedures established previously(8, 9). Reads were aligned to the human reference genome GRCh37 by BWA (version 0.7.12)(10). Picard (http://broadinstitute.github.io/picard/, version 1.129) was used for marking PCR duplication. Afterwards, the reads were realigned around potential indel regions by GATK IndelRealigner module (version 3.5) following the recommended procedures(11). The MuTect2 module from GATK was used to identify single nucleotide variants and indels from matched leukemia and germline samples(12). Variants with any of the following features in the tumor data were excluded: 1) read depth <20; 2) mutant allele frequency <10%; 3) all reads supporting mutation calls coming from the same mapping direction; and 4) two or more mutation called in the same sample within a 30 bp window. Remaining high quality variants were then annotated by ANNOVAR(13). Tumor copy-number variations and structural variations were detected using CONSERTING(14) and CREST(15).

Total RNA library was constructed using Illumina TrueSeq stranded mRNA library prep kit and sequenced using the HiSeq 2000/2500 or NovaSeq 6000 platform (2 x 101- bp pair-end reads). On average, we achieved at least 20x coverage for more than 30% of the transcriptome. Gene expression was quantified by STAR(16) (ver. 2.6.0b) under default parameters with the human genome (GRCh37) and annotation file (Gencode v19)(17, 18).

**Statistical analysis**

Statistical analysis was performed by Student t test or wilcoxon test. The p value of IP-MS data was derived by G-test (19).

**HDR template:**

Landing pad (EF1α-Bxb1_attP-BFP)

CCTTTGCAAGCAAACATCTTGACTGCTTTCTCTGACCAGCATTCTCTCCCCTGGGCCTGTGCCGCTTTCTGTCTGCAGCTTGTGGCCTGGGTCACCTCTACGGCTGGCCCAGATCCTTCCCTGCCGCCTCCTTCAGGTTCCGTCTTCCTCCACTCCCTCTTCCCCTTGCTCTCTGCTGTGTTGCTGCCCAAGGATGCTCTTTCCGGAGCACTTCCTTCTCGGCGCTGCACCACGTGATGTCCTCTGAGCGGATCCTCCCCGTGTCTGGGTCCTCTCCGGGCATCTCTCCTCCCTCACCCAACCCCATGCCGTCTTCACTCGCTGGGTTCCCTTTTCCTTCTCCTTCTGGGGCCTGTGCCATCTCTCGTTTCTTAGGATGGCCTTCTCCGACGGATGTCTCCCTTGCGTCCCGCCTCCCCTTCTTGTAGGCCTGCATCATCACCGTTTTTCTGGACAACCCCAAAGTACCCCGTCTCCCTGGCTTTAGCCACCTCTCCATCCTCTTGCTTTCTTTGCCTGGACACCCCGTTCTCCTGTGGATTCGGGTCACCTCTCACTCCTTTCATTTGGGCAGCTCCCCTACCCCCCTTACCTCTCTAGTCTGTGCTAGCTCTTCCAGCCCCCTGTCATGGCATCTTCCAGGGGTCCGAGAGCTCAGCTAGTCTTCTTCCTCCAACCCGGGCCCCTATGTCCACTTCAGGACAGCATGTTTGCTGCCTCCAGGGATCCTGTGTCCCCGAGCTGGGACCACCTTATATTCCCAGGGCCGGTTAATGTGGCTCTGGTTCTGGGTACTTTTATCTGTCCCCTCCACCCCACAGTGGGGCctaggtcttgaaaggagtgggaattggctccggtgcccgtcagtgggcagagcgcacatcgcccacagtccccgagaagttggggggaggggtcggcaattgaaccggtgcctagagaaggtggcgcggggtaaactgggaaagtgatgtcgtgtactggctccgcctttttcccgagggtgggggagaaccgtatataagtgcagtagtcgccgtgaacgttctttttcgcaacgggtttgccgccagaacacaggtaagtgccgtgtgtggttcccgcgggcctggcctctttacgggttatggcccttgcgtgccttgaattacttccacctggctgcagtacgtgattcttgatcccgagcttcgggttggaagtgggtgggagagttcgaggccttgcgcttaaggagccccttcgcctcgtgcttgagttgaggcctggcctgggcgctggggccgccgcgtgcgaatctggtggcaccttcgcgcctgtctcgctgcttt

cgataagtctctagccatttaaaatttttgatgacctgctgcgacgctttttttctggcaagatagtcttgtaaatgcgggccaagatctg
cacactggtatttcggtttttggggccgcgggcggcgacggggcccgtgcgtcccagcgcacatgttcggcgaggcggggcctg
cgagcgcggccaccgagaatcggacgggggtagtctcaagctggccggcctgctctggtgcctggcctcgcgccgccgtgtat
cgccccgccctgggcggcaaggctggcccggtcggcaccagttgcgtgagcggaaagatggccgcttcccggccctgctgca
gggagctcaaaatggaggacgcggcgctcgggagagcgggcgggtgagtcacccacacaaaggaaaagggcctttccgtc
ctcagccgtcgcttcatgtgactccacggagtaccgggcgccgtccaggcacctcgattagttctcgagcttttggagtacgtcgtct
ttaggttgggggggaggggttttatgcgatggagtttccccacactgagtgggtggagactgaagttaggccagcttggcacttgatg
taattctccttggaatttgccctttttgagtttggatcttggttcattctcaagcctcagacagtggttcaaagtttttttcttccatttcaggtgt
cgtgaGGAATTGATCCAGATCTGCTGGTTTGTCTGGTCAACCACCGCGGTCTCAGTGGTG
TACGGTACAAACCGCCACCATGGTGTCTAAGGGCGAAGAGCTGATTAAGGAGAACATGC
ACATGAAGCTGTACATGGAGGGCACCGTGGACAACCATCACTTCAAGTGCACATCCGAG
GGCGAAGGCAAGCCCTACGAGGGCACCCAGACCATGAGAATCAAGGTGGTCGAGGGC
GGCCCTCTCCCCTTCGCCTTCGACATCCTGGCTACTAGCTTCCTCTACGGCAGCAAGAC
CTTCATCAACCACACCCAGGGCATCCCCGACTTCTTCAAGCAGTCCTTCCCTGAGGGCT
TCACATGGGAGAGAGTCACCACATACGAAGACGGGGGCGTGCTGACCGCTACCCAGGA
CACCAGCCTCCAGGACGGCTGCCTCATCTACAACGTCAAGATCAGAGGGGTGAACTTC
ACATCCAACGGCCCTGTGATGCAGAAGAAAACACTCGGCTGGGAGGCCTTCACCGAGA
CGCTGTACCCCGCTGACGGCGGCCTGGAAGGCAGAAACGACATGGCCCTGAAGCTCG
TGGGCGGGAGCCATCTGATCGCAAACGCCAAGACCACATATAGATCCAAGAAACCCGCT
AAGAACCTCAAGATGCCTGGCGTCTACTATGTGGACTACAGACTGGAAAGAATCAAGGA
GGCCAACAACGAGACCTACGTCGAGCAGCACGAGGTGGCAGTGGCCAGATACTGCGA
CCTCCCTAGCAAACTGGGGCACAAGCTTAATTAAGGATCCATCGGATCCCGGGCCCGTC

GACGGTACCCTGTGCCTTCTAGTTGCCAGCCATCTGTTGTTTGCCCCTCCCCCGTGCCT
TCCTTGACCCTGGAAGGTGCCACTCCCACTGTCCTTTCCTAATAAAATGAGGAAATTGCA
TCGCATTGTCTGAGTAGGTGTCATTCTATTCTGGGGGGTGGGGTGGGGCAGGACAGCA
AGGGGGAGGATTGGGAAGACAATAGCAGGCATGCTGGGGATGCGGTGGGCTCTATGGT
TACTAGGGACAGGATTGGTGACAGAAAAGCCCCATCCTTAGGCCTCCTCCTTCCTAGTC
TCCTGATATTGGGTCTAACCCCCACCTCCTGTTAGGCAGATTCCTTATCTGGTGACACAC
CCCCATTTCCTGGAGCCATCTCTCTCCTTGCCAGAACCTCTAAGGTTTGCTTACGATGGA
GCCAGAGAGGATCCTGGGAGGGAGAGCTTGGCAGGGGGTGGGAGGGAAGGGGGGGA
TGCGTGACCTGCCCGGTTCTCAGTGGCCACCCTGCGCTACCCTCTCCCAGAACCTGAG
CTGCTCTGACGCGGCTGTCTGGTGCGTTTCACTGATCCTGGTGCTGCAGCTTCCTTACA
CTTCCCAAGAGGAGAAGCAGTTTGGAAAAACAAAATCAGAATAAGTTGGTCCTGAGTTC
TAACTTTGGCTCTTCACCTTTCTAGTCCCCAATTTATATTGTTCCTCCGTGCGTCAGTTTT
ACCTGTGAGATAAGGCCAGTAGCCAGCCCCGTCCTGGCAGGGCTGTGGTGAGGAGGG
GGGTGTCCGTGTGGAAAACTCCCTTTGTGAGAATGGTGCGTCCTAGGTGTTCACCAGG
TCGTGGCCGCCTCTACTCCCTTTCTCTTTCTCCATCCTTCTTTCCTTAAAGAGTCCCCAG
TGCTATCTGGGACATATTCCTCCGCCCAGAGCAGGGTCCCGCTTCCCTAAGGCCCTGCT
CTGGGCTTCTGGGTTTGAGTCCTTGGCAAGCCCAGGAGAGGCGCTCAGGCTTCCCTGT
CCCCCTTCCTCGTCCACCATCTCATGCCCCTGGCTCTCCTGCCCCTTCCCTACAGGGGT
TCCTGGCTCTGCTCTGTCAAGATGTTTGCTTGCAAAGG

GZMA-TY1-P2A-EGFP

CCTTTGCAAGCAAACATCTTGACTGCCTGAAAGGGACTGATTTGGTTTTGTTTCTTTTGG
AAGGCAATTATCTGCTAGAAGAACCACAAAACATAGTGTTTATTCTTTGCTTCAATGTATCA

TCTGCATTTGACTATTTTGCCCCTTGAGTTATTAAGCATTTTGAGAAAACGACAAATAAAC

AGGAGACTTTCCTTTCCAAAAGCAGAGCAATAGTCTCAAAATTAGCTGATAACATGTACAA

GTTTCACTTCGATTTTCACTGGCTCATAAATAAACCAAGTGAACCAATTCAAAAATATTAAA

ATATTTCCAAACATTTTAATTTTTAAAATTAAAGCACTATCTTCAATTAAGTCAAGGTTGGTC

TTAACTGCATATTAAATGCTGCAGAATTTCTTCCATTTCACTAGTGGTAATGCTGAACACT

GACCCCACACCCTACCCCTCTTGTTTTCCTCCAGGGAGATTCTGGAAGCCCTTTGTTGT

GCGAGGGTGTTTTCCGAGGGGTCACTTCCTTTGGCCTTGAAAATAAATGCGGAGACCCT

CGTGGGCCTGGTGTCTATATTCTTCTCTCAAAGAAACACCTCAACTGGATAATTATGACTA

TCAAGGGAGCAGTTGAGGTGCACACCAACCAGGACCCCCTGGACGCCGAAGTCCATAC

AAATCAGGATCCTCTGGATGCCGAAGTGCACACCAATCAGGATCCCCTGGACGCTGGAA

GCGGAGCTACTAACTTCTCTCTGTTAAAGCAAGCAGGAGACGTGGAAGAAAACCCCGG

TCCCATGGTGAGCAAGGGCGAGGAGCTGTTCACCGGGGTGGTGCCCATCCTGGTCGA

GCTGGACGGCGACGTAAACGGCCACAAGTTCAGCGTGTCCGGCGAGGGCGAGGGCG

ATGCCACCTACGGCAAGCTGACCCTGAAGTTCATCTGCACCACCGGCAAGCTGCCCGT

GCCCTGGCCCACCCTCGTGACCACCCTGACCTACGGCGTGCAGTGCTTCAGCCGCTAC

CCCGACCACATGAAGCAGCACGACTTCTTCAAGTCCGCCATGCCCGAAGGCTACGTCC

AGGAGCGCACCATCTTCTTCAAGGACGACGGCAACTACAAGACCCGCGCCGAGGTGAA

GTTCGAGGGCGACACCCTGGTGAACCGCATCGAGCTGAAGGGCATCGACTTCAAGGA

GGACGGCAACATCCTGGGGCACAAGCTGGAGTACAACTACAACAGCCACAACGTCTATA

TCATGGCCGACAAGCAGAAGAACGGCATCAAGGTGAACTTCAAGATCCGCCACAACATC

GAGGACGGCAGCGTGCAGCTCGCCGACCACTACCAGCAGAACACCCCCATCGGCGAC

GGCCCCGTGCTGCTGCCCGACAACCACTACCTGAGCACCCAGTCCGCCCTGAGCAAA

GACCCCAACGAGAAGCGCGATCACATGGTCCTGCTGGAGTTCGTGACCGCCGCCGGG
ATCACTCTCGGCATGGACGAGCTGTACAAGTGAGGATCCTGATCATAATCAGCCATACCA
CATTTGTAGAGGTTTTACTTGCTTTAAAAAACCTCCCACACCTCCCCCTGAACCTGAAAC
ATAAAATGAATGCAATTGTTGTTGTTAACTTGTTTATTGCAGCTTATAATGGTTACAAATAAA
GCAATAGCATCACAAATTTCACAAATAAAGCATTTTTTTCACTGCATTCTAGTTGTGGTTTG
TCCAAACTCATCAATGTATCTTATAAATAACCGTTTgCTTTCATTTACTGTGGCTTCTTAATC
TTTTCACAAATAAAATCAATTTGCATGACTGTACCTGTTTCTCTCTTGTAACCTTAGTGGG
CAGATCTGCACCAGCAAAGTGAAGCAGAGTTACATGGCAGCCTTGTAGATAATGCAAGG
ATTACGTGATCAATGTCACCAGAGTCATTTGTGCGTCAAGTGACATGGGAATGCTTCCTG
AATTATTCATCTCTCTGTCTCCATGAAATCCAGCAAGAACAACCCTAAGATCAGGTCTAAT
AACAAATGCAGTAAAGGCCTCCACAATTCCACAGGAAACAGAAACAGCCAGTCTGGAAC
ATTTGTGTTAAAAGGAAATATAGGGCATTGTTCCACATTAACACCTGCTTTTCATGTTATAC
ACAAGAACCTGAGTCTATGGGAGAAAAAGAAACAAGCAATGCCTTACGGTATTTTCCAAA
TTCTAGGGCATACAAATAAAGTGCTTGGGCAGCCAAGAAAATTATACAATAGAGAGTCAG
CCTTCTTTCTCAGTGCCTTGCATTTAAGCCTTGGATTACCTAGGTGATTTCTCAGTTCTCT
TTGTTCTTGAGACGGGGGTCAAGATGTTTGCTTGCAAAGG
p.G365R-MUT-LHA-3HA-P2A-mCherry-RHA
CCTTTGCAAGCAAACATCTTGACCATCCTGAGTGGTCCCCGACCTCCTGGGCATAGCAT
CATGGGTAGTCCCCATCCTCTTGGGAGGTGACATGCTGGGTGATCCTCGTCATCTCAGG
AGGTGGCATCCTGGGTGGTCCCTGTCCCCCTGGGTATAGCATCCTGGGTAATCCTCGTC
CTCTTGGGAGTAGCATCCCGGGTGGTCCCCGTCCTCCCCAGCAGTAGCATCCTGGGTG
GCTTCCCATCCTCCTAGGCGGTATCATCCTGGGTAGCCCCCTGGGGCAGAGGGAAGAG

CTGTGGCCTCCGCAACCTCCTACTCACTTCCGCTCCGTTCTCTTGCCCGCCCTGCAGCGGCACCCGACCTGACAGCGTTCAGCGACCCGCGCCAGTTCCCCGCGCTGCCCTCCATCTCCGACCCCCGCATGCACTATCCAGGCGCCTTCACCTACTCCCCGACGCCGGTCACCTCGGGCATCCGCATTGGTATGTCCGCGATGGGGTCCGCGACCCGGTATCATACGTATCTCCCCCCGCCGTATCCGGGGTCCTCCCAGGCCCAAGGTGGGCCCTTTCAGGCGAGTTCCCCGTCGTATCATCTATATTATGGGGCGTCCGCGGGGTCGTATCAATTTTCGATGGTAGGGGGCGAACGGTCCCCCCCGCGGATTCTACCGCCGTGTACGAATGCGTCGACGGGGTCGGCCCTACTTAATCCGAGTCTTCCCAATCAAAGTGATGTAGTAGAAGCGGAAGGGAGTCATAGTAATTCTCCGACGAATATGGCGCCGTCGGCGCGGCTAGAAGAAGCGGTATGGAGACCGTATTACCCATACGATGTTCCTGACTATGCGGGCTATCCCTATGACGTCCCGGACTATGCAGGATCCTATCCATATGACGTTCCAGATTACGCTGGAAGCGGAGCTACTAACTTCTCTCTGTTAAAGCAAGCAGGAGACGTGGAAGAAAACCCCGGTCCCGTGAGCAAGGGCGAGGAGGATAACATGGCCATCATCAAGGAGTTCATGCGCTTCAAGGTGCACATGGAGGGCTCCGTGAACGGCCACGAGTTCGAGATCGAGGGCGAGGGCGAGGGCCGCCCCTACGAGGGCACCCAGACCGCCAAGCTGAAGGTGACCAAGGGTGGCCCCCTGCCCTTCGCCTGGGACATCCTGTCCCCTCAGTTCATGTACGGCTCCAAGGCCTACGTGAAGCACCCCGCCGACATCCCCGACTACTTGAAGCTGTCCTTCCCCGAGGGCTTCAAGTGGGAGCGCGTGATGAACTTCGAGGACGGCGGCGTGGTGACCGTGACCCAGGACTCCTCCCTGCAGGACGGCGAGTTCATCTACAAGGTGAAGCTGCGCGGCACCAACTTCCCCTCCGACGGCCCCGTAATGCAGAAGAAGACCATGGGCTGGGAGGCCTCCTCCGAGCGGATGTACCCCGAGGACGGCGCCCTGAAGGGCGAGATCAAGCAGAGGCTGAAGCTGAAGGACGGCGGCCACTACGACGCTGAGGTCAAGACCACCTACAAGGCCAAGAAGCCCGTGCAGCTGCCCGGCGCCT

ACAACGTCAACATCAAGTTGGACATCACCTCCCACAACGAGGACTACACCATCGTGGAACAGTACGAACGCGCCGAGGGCCGCCACTCCACCGGCGGCATGGACGAGCTGTACAAGTGATGCACCAGCCCTGGCCCGGCTGGGCCCCGCGGGCCGCCGCCTTCGCCTCCGGGCGCGCGGGCCTCCTGTTCGCGACAAGCCCGCCGGGATCCCGGGCCCTGGGCCCGGCCACCGTCCTGGGGCCGAGGGCGCCCGACGGCCAGGATCTCGCTGTAGGTCAGGCCCGCGCAGCCTCCTGCGCCCAGAAGCCCACGCCGCCGCCGTCTGCTGGCGCCCCGGCCCTCGCGGAGGTGTCCGAGGCGACGCACCTCGAGGGTGTCCGCCGGCCCCAGCACCCAGGGGACGCGCTGGAAAGCAAACAGGAAGATTCCCGGAGGGAAACTGTGAATGCTTCTGATTTAGCAATGCTGTGAATAAAAAGAAAGATTTTATACCCTTGACTTAACTTTTTAACCAAGTTGTTTATTCCAAAGAGTGTGGAATTTTGGTTGGGGTGGGGGGAGAGGAGGGGTCAAGATGTTTGCTTGCAAAGG

p.G365R-WT-LHA-TY1-P2A-EGFP-RHA

CCTTTGCAAGCAAACATCTTGACCATCCTGAGTGGTCCCCGACCTCCTGGGCATAGCATCATGGGTAGTCCCCATCCTCTTGGGAGGTGACATGCTGGGTGATCCTCGTCATCTCAGGAGGTGGCATCCTGGGTGGTCCCTGTCCCCCTGGGTATAGCATCCTGGGTAATCCTCGTCCTCTTGGGAGTAGCATCCCGGGTGGTCCCCGTCCTCCCCAGCAGTAGCATCCTGGGTGGCTTCCCATCCTCCTAGGCGGTATCATCCTGGGTAGCCCCCTGGGGCAGAGGGAAGAGCTGTGGCCTCCGCAACCTCCTACTCACTTCCGCTCCGTTCTCTTGCCCGCCCTGCAGcggcacccgacctgacagcgttcagcgacccgcgccagttccccgcgctgccctccatctccgacccccgcatgcactatccaggcgccttcacctactccccgacgccggtcacctcgggcatcggcatcggcatgtcggccatgggctcggccacgcgctaccacacctacctgccgccgccctaccccggctcgtcgcaagcgcagggaggcccgttccaagccagctcgccctcctaccacctgtactacggcgcctcggccggctcctaccagttctccatggtgggcggcgagcgctcgccgccgcgcatcctgccgccctgcaccaacg cctccaccggctccgcgctgctcaaccccagcctcccgaaccagagcgacgtggtggaggccgagggcagccacagcaactcccccaccaacatggcgccctccgcgcgcctggaggaggccgtgtggaggccctacGAGGTGCACACCAACCAGGACCCCCTGGACGCCGAAGTCCATACAAATCAGGATCCTCTGGATGCCGAAGTGCACACCAATCAGGATCCCCTGGACGCTGGAAGCGGAGCTACTAACTTCTCTCTGTTAAAGCAAGCAGGAGACGTGGAAGAAAACCCCGGTCCCATGGTGAGCAAGGGCGAGGAGCTGTTCACCGGGGTGGTGCCCATCCTGGTCGAGCTGGACGGCGACGTAAACGGCCACAAGTTCAGCGTGTCCGGCGAGGGCGAGGGCGATGCCACCTACGGCAAGCTGACCCTGAAGTTCATCTGCACCACCGGCAAGCTGCCCGTGCCCTGGCCCACCCTCGTGACCACCCTGACCTACGGCGTGCAGTGCTTCAGCCGCTACCCCGACCACATGAAGCAGCACGACTTCTTCAAGTCCGCCATGCCCGAAGGCTACGTCCAGGAGCGCACCATCTTCTTCAAGGACGACGGCAACTACAAGACCCGCGCCGAGGTGAAGTTCGAGGGCGACACCCTGGTGAACCGCATCGAGCTGAAGGGCATCGACTTCAAGGAGGACGGCAACATCCTGGGGCACAAGCTGGAGTACAACTACAACAGCCACAACGTCTATATCATGGCCGACAAGCAGAAGAACGGCATCAAGGTGAACTTCAAGATCCGCCACAACATCGAGGACGGCAGCGTGCAGCTCGCCGACCACTACCAGCAGAACACCCCCATCGGCGACGGCCCCGTGCTGCTGCCCGACAACCACTACCTGAGCACCCAGTCCGCCCTGAGCAAAGACCCCAACGAGAAGCGCGATCACATGGTCCTGCTGGAGTTCGTGACCGCCGCCGGGATCACTCTCGGCATGGACGAGCTGTACAAGtgaTgcAccagCcctggcccggctgggccccgcgggccgccgccttcgcctccgggcgcgcgggcctcctgttcgcgacaagcccgccgggatcccgggccctgggcccggccaccgtcctggggccgagggcgcccgacggccaggatctcgctgtaggtcaggcccgcgcagcctcctgcgcccagaagcccacgccgccgccgtctgctggcgccccggccctcgcggaggtgtccgaggcgacgcacctcgagggtgtccgccggccccagcacccaggggacgcgctggaaagcaaacaggaagattcccggagggaaactgtgaatgcttctgatttagcaatgctgtgaataaaaagaaagattttataccсttgacttaactttttaaccaagttgtttattccaaaga gtgtggaattttggttggggtggggggagaggagggatgcaactcgccctgtttggcatctaattcttatttttaatttttccgcaccttatcaattgcaaaatgcgtatttgcatttgggtggtttttatttttatatacgtttatataaatatatataaattgagcttgcttctttcttgctttgaccatggaaagaaatatgattcccttttctttaagttttatttaacttttcttttggacttttgggtagttgttttttttgttttgttttgtttttttgagaaacagctacagctttgggtcatttttaactactgtattcccacaaggaatccccagatatttatgtatcttgatgtTCAGACATTTATGTGTTGTCAAGATGTTTGCTTGCAAAGG

p.Y287*-MUT-LHA-3HA-P2A-mCherry-RHA

CCTTTGCAAGCAAACATCTTGACAGACTTTTGTCTCTTCTCTATCCCAGGCCTCTTATAATGGCAATCTATAAAATACCTGCTAAAAACTCTGACTTTTAAAAGTTGCAAACTCGAAGTCCTGAAAAGGAACAGAAGGCTTGGGTTGATAGGGCCAGTTCTACTGCTGGGTTTGTGAACTAGTCATTAGACCTCATTGCCGGTTTCCCAGCCAGACCTTCTTTCCTGTCATCCTTGGGAGAGAATTCGCCTTACTATAAAACATTTACCAGCCCATGGAATACTTTTTGCCAAAGGTAAAGGTATAAAAAAAAAGCCCAAAACTCTACGTTCTTACCGAACACAGACAGGTGATCCCCTGAGTTTATAAAATGTTTCTGATGATGATAATTGTAATTATTGGCAAAGCTGAAATCATGTCAGCAGTCATATGATACTTGAAGTTCTCAAGTGCTGCAGTTCAACAGTCTGGACATTAATACTTCTACCTTGAAAGTGTAATCCATTCCTCTCATCATTGTTAGCATGATTAAATAAAACCTTTGGAAGGAATAGTTATCAGGTGAAAATCTCCAAGAATCAGTCTCTTTTGGGGGAAAATAATCCAACAGAGGCAGATACTTGGACTTGAGTAGGCTTATTAAACCCTGGTACATAGGCCACATACATGTATGTGACATATTTGAACAAGGGCCACTCATTTCTTATTAAAAGACATTTTTTAAATCCCACCCCACTTTACATATAATTGACCTTTCTGATTCTCTTCAGatacaaggcagatccaaccatccccaccgtggtcctacgatcagtcctaTcaaTACCCATACGATGTTCCTGACTATGCGGGCTATCCCTATGACGTCCCGGACTATGCAGGATCCTATCCATATGACGTTCCAGATTACGCTGGAAGCGGAGCTACTAACTTCTCTCTGTTAAAGCAAGCAGGAGACGTGGAAGAAAACCCCGGTCCCGTGA

GCAAGGGCGAGGAGGATAACATGGCCATCATCAAGGAGTTCATGCGCTTCAAGGTGCACATGGAGGGCTCCGTGAACGGCCACGAGTTCGAGATCGAGGGCGAGGGCGAGGGCCGCCCCTACGAGGGCACCCAGACCGCCAAGCTGAAGGTGACCAAGGGTGGCCCCCTGCCCTTCGCCTGGGACATCCTGTCCCCTCAGTTCATGTACGGCTCCAAGGCCTACGTGAAGCACCCCGCCGACATCCCCGACTACTTGAAGCTGTCCTTCCCCGAGGGCTTCAAGTGGGAGCGCGTGATGAACTTCGAGGACGGCGGCGTGGTGACCGTGACCCAGGACTCCTCCCTGCAGGACGGCGAGTTCATCTACAAGGTGAAGCTGCGCGGCACCAACTTCCCCTCCGACGGCCCCGTAATGCAGAAGAAGACCATGGGCTGGGAGGCCTCCTCCGAGCGGATGTACCCCGAGGACGGCGCCCTGAAGGGCGAGATCAAGCAGAGGCTGAAGCTGAAGGACGGCGGCCACTACGACGCTGAGGTCAAGACCACCTACAAGGCCAAGAAGCCCGTGCAGCTGCCCGGCGCCTACAACGTCAACATCAAGTTGGACATCACCTCCCACAACGAGGACTACACCATCGTGGAACAGTACGAACGCGCCGAGGGCCGCCACTCCACCGGCGGCATGGACGAGCTGTACAAGtagGGATCCTGATCATAATCAGCCATACCACATTTGTAGAGGTTTTACTTGCTTTAAAAAACCTCCCACACCTCCCCCTGAACCTGAAACATAAAATGAATGCAATTGTTGTTGTTAACTTGTTTATTGCAGCTTATAATGGTTACAAATAAAGCAATAGCATCACAAATTTCACAAATAAAGCATTTTTTTCACTGCATTCTAGTTGTGGTTTGTCCAAACTCATCAATGTATCTTActgggatccattgcctctccttctgtgcacccagcaacgcccatttcacctggacgtgccagcggcatgacaaccctctctgcagaactttccagtcgactctcaaGTAAGCCACTTGAAAACACATTCTTTGCAGCTGAGCTGGGGTGGAAGGTCCAGGAGACTAGAGGTGCATGAAGGAGTTGGCAGAACATTATTGAGTAAAACATGGTTTAAAGGTGACCTATCTCTACTCTGAGTCCCATGCCCATGCACTGATTTTCTTGGATACATAAGAGGCAGAGATTGTGGATGTGAAGGCAGAAGTGGCTTGGAGAAGTAGGTTCCTTAGACATCTAAAGTTGTTCACATTTGGCAAGAAAGGATGGTGACTGAAGGGTTAAGAA

CAATATAAAAAGCCAAAGTTTTCAACTTTTAGGACTCATTATTCCAAATGAATTCTGGACTAATGGGACTGGTCCATGCGGTGGTGGTAAATTTTCACAGATCACATGGACAGCACACGGGAGTACATGTGGAATGGGAAATAAATACAGCCGCCCCTTGGTATCCATGGAGGACTGGTTCCAGGACCCCCCATGGATACCAAAACCCATGGATGCTCAAGTCCCTCATATAAAATGGTATATTTATATATAACATATATATCCTCCCATGTATTTAAATCATCCCTAGATTACTCGTAATACCTAATAAAATGTAAATGCTATGTAAATACTTGTTATACTGCATTGTTTAGGGAATGACAAGAAGAAAAGTCTGTATATGTCTAATACAGACACAACCATCCTTTTTTTGTCAAGATGTTTGCTTGCAAAGG

p.Y287*-WT-LHA-TY1-P2A-EGFP-RHA

CCTTTGCAAGCAAACATCTTGACAGACTTTTGTCTCTTCTCTATCCCAGGCCTCTTATAATGGCAATCTATAAAATACCTGCTAAAAACTCTGACTTTTAAAAGTTGCAAACTCGAAGTCCTGAAAAGGAACAGAAGGCTTGGGTTGATAGGGCCAGTTCTACTGCTGGGTTTGTGAACTAGTCATTAGACCTCATTGCCGGTTTCCCAGCCAGACCTTCTTTCCTGTCATCCTTGGGAGAGAATTCGCCTTACTATAAAACATTTACCAGCCCATGGAATACTTTTTGCCAAAGGTAAAGGTATAAAAAAAAAGCCCAAAACTCTACGTTCTTACCGAACACAGACAGGTGATCCCCTGAGTTTATAAAATGTTTCTGATGATGATAATTGTAATTATTGGCAAAGCTGAAATCATGTCAGCAGTCATATGATACTTGAAGTTCTCAAGTGCTGCAGTTCAACAGTCTGGACATTAATACTTCTACCTTGAAAGTGTAATCCATTCCTCTCATCATTGTTAGCATGATTAAATAAAACCTTTGGAAGGAATAGTTATCAGGTGAAAATCTCCAAGAATCAGTCTCTTTTGGGGGAAAATAATCCAACAGAGGCAGATACTTGGACTTGAGTAGGCTTATTAAACCCTGGTACATAGGCCACATACATGTATGTGACATATTTGAACAAGGGCCACTCATTTCTTATTAAAAGACATTTTTTAAATCCCACCCCACTTTACATATAATTGACCTTTCTGATTCTCTTCAGatacaaggcagatccaaccatcccca

ccgtggtcctacgatcagtcctaTcaataTctTggGtcTatCgcAtcCccAtcCgtTcaTccTgcCacCccTatCtcGccA
ggGcgCgcGagTggTatgacTacGctTtcCgcCgaGctCtcTagCcgGctTtcGacggcacccgacctgacagcgtt
cagcgacccgcgccagttccccgcgctgccctccatctccgaccccgcatgcactatccaggcgccttcacctactccccgac
gccggtcacctcgggcatcggcatcggcatgtcggccatgggctcggccacgcgctaccacacctacctgccgccgccctacc
ccggctcgtcgcaagcgcagggaggcccgttccaagccagctcgccctcctaccacctgtactacggcgcctcggccggctcct
accagttctccatggtgggcggcgagcgctcgccgccgcgcatcctgccgccctgcaccaacgcctccaccggctccgcgctg
ctcaaccccagcctcccgaaccagagcgacgtggtggaggccgagggcagccacagcaactcccccaccaacatggcgcc
ctccgcgcgcctggaggaggccgtgtggaggccctacGAGGTGCACACCAACCAGGACCCCCTGGACG
CCGAAGTCCATACAAATCAGGATCCTCTGGATGCCGAAGTGCACACCAATCAGGATCCC
CTGGACGCTGGAAGCGGAGCTACTAACTTCTCTCTGTTAAAGCAAGCAGGAGACGTGG
AAGAAAACCCCGGTCCCATGGTGAGCAAGGGCGAGGAGCTGTTCACCGGGGTGGTGC
CCATCCTGGTCGAGCTGGACGGCGACGTAAACGGCCACAAGTTCAGCGTGTCCGGCG
AGGGCGAGGGCGATGCCACCTACGGCAAGCTGACCCTGAAGTTCATCTGCACCACCG
GCAAGCTGCCCGTGCCCTGGCCCACCCTCGTGACCACCCTGACCTACGGCGTGCAGT
GCTTCAGCCGCTACCCCGACCACATGAAGCAGCACGACTTCTTCAAGTCCGCCATGCC
CGAAGGCTACGTCCAGGAGCGCACCATCTTCTTCAAGGACGACGGCAACTACAAGACC
CGCGCCGAGGTGAAGTTCGAGGGCGACACCCTGGTGAACCGCATCGAGCTGAAGGGC
ATCGACTTCAAGGAGGACGGCAACATCCTGGGGCACAAGCTGGAGTACAACTACAACA
GCCACAACGTCTATATCATGGCCGACAAGCAGAAGAACGGCATCAAGGTGAACTTCAAG
ATCCGCCACAACATCGAGGACGGCAGCGTGCAGCTCGCCGACCACTACCAGCAGAACA
CCCCCATCGGCGACGGCCCCGTGCTGCTGCCCGACAACCACTACCTGAGCACCCAGT
CCGCCCTGAGCAAAGACCCCAACGAGAAGCGCGATCACATGGTCCTGCTGGAGTTCGT

GACCGCCGCCGGGATCACTCTCGGCATGGACGAGCTGTACAAGtgaGGATCCTGATCAT
AATCAGCCATACCACATTTGTAGAGGTTTTACTTGCTTTAAAAAACCTCCCACACCTCCCC
CTGAACCTGAAACATAAAATGAATGCAATTGTTGTTGTTAACTTGTTTATTGCAGCTTATAA
TGGTTACAAATAAAGCAATAGCATCACAAATTTCACAAATAAAGCATTTTTTTCACTGCATT
CTAGTTGTGGTTTGTCCAAACTCATCAATGTATCTTActgggatccattgcctctccttctgtgcacccagca
acgcccatttcacctggacgtgccagcggcatgacaaccctctctgcagaactttccagtcgactctcaaGTAAGCCACTT
GAAAACACATTCTTTGCAGCTGAGCTGGGGTGGAAGGTCCAGGAGACTAGAGGTGCAT
GAAGGAGTTGGCAGAACATTATTGAGTAAAACATGGTTTAAAGGTGACCTATCTCTACTC
TGAGTCCCATGCCCATGCACTGATTTTCTTGGATACATAAGAGGCAGAGATTGTGGATGT
GAAGGCAGAAGTGGCTTGGAGAAGTAGGTTCCTTAGACATCTAAAGTTGTTCACATTTG
GCAAGAAAGGATGGTGACTGAAGGGTTAAGAACAATATAAAAAGCCAAAGTTTTCAACTT
TTAGGACTCATTATTCCAAATGAATTCTGGACTAATGGGACTGGTCCATGCGGTGGTGGT
AAATTTTCACAGATCACATGGACAGCACACGGGAGTACATGTGGAATGGGAAATAAATAC
AGCCGCCCCTTGGTATCCATGGAGGACTGGTTCCAGGACCCCCCATGGATACCAAAAC
CCATGGATGCTCAAGTCCCTCATATAAAATGGTATATTTATATATAACATATATATCCTCCCAT
GTATTTAAATCATCCCTAGATTACTCGTAATACCTAATAAAATGTAAATGCTATGTAAATACTT
GTTATACTGCATTGTTTAGGGAATGACAAGAAGAAAAGTCTGTATATGTCTAATACAGACA
CAACCATCCTTTTTTTGTCAAGATGTTTGCTTGCAAAGG

p.R232fs-MUT-LHA-3HA-P2A-mCherry-RHA

cctttgcaagcaaacatcttgactcctgtcaactcctccttaggggtacatgggggagtcggcttcagggaagaaggaatgcttgt
gaaaagcctgtgaggagagggagagggagaggggctgaggggctgaggatgccctgcgtgtacctggaaatgggacctcc
ccagaggctgtgggtttgcagccacgcctcatttcctcttggaaggggccctaccactaagctgcggggcccactttgccgcctct

aaaacgtgccagtggctgggtttgcagtaaggaacagccgttctgatgcccctgggaggccacctttcctggctgccacagcata

gcccctggcatgtggccatgtgccaacaattaatgcgcctctttccctcccggcatcaccaccaccgccaagttctgtatctcagtg

agttgtgcccattggaagtgggtgtagagggttggggagggtgccaaggcccagactgggggagcactctgtggccgaggcg

gtgaaaggggggcccattctgctgagaggacagtggccccaaattcagctggcatatctctagcgagtctatgttggggtgaggg

gagagagaggggaaagacaagaaaagccccagttttaggaaatccacaatactttttctgatctcttccctccctccttccctccc

cccatcccctcccctccctgctccccacaataggacatcggcagaaactagatgatcagaccaagcccgggagcttgtccttttc

cgagcggctcagtgaactcgagcagctgcgcgcacagccatacccatacgatgttcctgactatgcgggctatccctatgacgtc

ccggactatgcaggatcctatccatatgacgttccagattacgctggaagcggagctactaacttctctctgttaaagcaagcagg

agacgtggaagaaaaccccggtcccgtgagcaagggcgaggaggataacatggccatcatcaaggagttcatgcgcttcaa

ggtgcacatggagggctccgtgaacggccacgagttcgagatcgagggcgagggcgagggccgcccctacgagggcaccc

agaccgccaagctgaaggtgaccaagggtggcccccctgcccttcgcctgggacatcctgtcccctcagttcatgtacggctcca

aggcctacgtgaagcaccccgccgacatccccgactacttgaagctgtccttccccgagggcttcaagtgggagcgcgtgatga

acttcgaggacggcggcgtggtgaccgtgacccaggactcctccctgcaggacggcgagttcatctacaaggtgaagctgcgc

ggcaccaacttcccctccgacggccccgtaatgcagaagaagaccatgggctgggaggcctcctccgagcggatgtaccccg

aggacggcgccctgaagggcgagatcaagcagaggctgaagctgaaggacggcggccactacgacgctgaggtcaagac

cacctacaaggccaagaagcccgtgcagctgcccggcgcctacaacgtcaacatcaagttggacatcacctcccacaacga

ggactacaccatcgtggaacagtacgaacgcgccgagggccgccactccaccggcggcatggacgagctgtacaagtgag

gatcctgatcataatcagccataccacatttgtagaggttttacttgctttaaaaaacctcccacacctccccctgaacctgaaacat

aaaatgaatgcaattgttgttgttaacttgtttattgcagcttataatggttacaaataaagcaatagcatcacaaatttcacaaataa

agcatttttttcactgcattctagttgtggtttgtccaaactcatcaatgtatcttagggtcagcccacaccacccagcccccacgccc

aaccctcgtgcctccctgaaccactccactgcctttaaccctcagcctcagagtcagatgcagggtaagtaccagatggagccc

actgcccgcctctcctgcacctgggccaccacccacaactggcccccatgtgcacacaccttcccagaccaactggggtttccc

cttgatgctcagagaaaggcctcgaaccaacagcaccacctgggagctgttggaaatgcagagtcttgggccccacccacccc
agacccaccgagtcagatctgcattgtaaccagatccccacaaggaaaagcactgctacagaggatacgaggagctctgggt
atggatatccatattggatagatagctctcttgaatttttaagagttagctttggctatgctatttttttacctacccattaggcttgaagac
acacacgcagacacacacacacacacacgtcaagcaaagtgaagatgggatgtttcaagttctcgctattgccagattatttgtg
ggttttgtatctagtgtttttttattattaagaaatagttgaaatgtatggatgtcatcacatcaagggtgttatttgcaaatcaatagaga
atgcaggtccccccagcccatggggctagctggcaattactaaagcgctgtaagatgcaataattgcctaaggcccactgtgcc
aaattagataatacaagaagttcatttacactgtagaccagtgacgtcaatgactgtttgctctgtgataccgtttcgtcaagatgtttg
cttgcaaagg

p.R232fs-WT-LHA-TY1-P2A-EGFP-RHA

CCTTTGCAAGCAAACATCTTGACTCCTGTCAACTCCTCCTTAGGGGTACATGGGGGAGT
CGGCTTCAGGGAAGAAGGAATGCTTGTGAAAAGCCTGTGAGGAGAGGGAGAGGGAGA
GGGGCTGAGGGGCTGAGGATGCCCTGCGTGTACCTGGAAATGGGACCTCCCCAGAGG
CTGTGGGTTTGCAGCCACGCCTCATTTCCTCTTGGAAGGGGCCCTACCACTAAGCTGC
GGGGCCCACTTTGCCGCCTCTAAAACGTGCCAGTGGCTGGGTTTGCAGTAAGGAACAG
CCGTTCTGATGCCCCTGGGAGGCCACCTTTCCTGGCTGCCACAGCATAGCCCCTGGCA
TGTGGCCATGTGCCAACAATTAATGCGCCTCTTTCCCTCCCGGCATCACCACCACCGCC
AAGTTCTGTATCTCAGTGAGTTGTGCCCATTGGAAGTGGGTGTAGAGGGTTGGGGAGG
GTGCCAAGGCCCAGACTGGGGGAGCACTCTGTGGCCGAGGCGGTGAAAGGGGGCCC
ATTCTGCTGAGAGGACAGTGGCCCCAAATTCAGCTGGCATATCTCTAGCGAGTCTATGTT
GGGGTGAGGGGAGAGAGAGGGGAAAGACAAGAAAAGCCCCAGTTTTAGGAAATCCAC
AATACTTTTTCTGATCTCTTCCCTCCCTCCTTCCCTCCCCCCATCCCCTCCCCTCCCTGC
TCCCCACAATAGGACATCGGCAGAAACTAGATGATCAGACCAAGCCCGGGAGCTTGTCC

TTTTCCGAGCGGCTCAGTGAACTCGAGCAGCTGCGACGCACAGCCATGAGAGTTTCCC
CTCATCATCCTGCGCCTACCCCTAATCCACGCGCATCTTTAAATCActccactgcctttaacccAC
AACCACAATCTCAAATGCAAGATACAAGGCAGATCCAACCATCCCCACCGTGGTCCTAC
GATCAGTCCTACCAATACCTGGGATCCATTGCCTCTCCTTCTGTGCACCCAGCAACGCC
CATTTCACCTGGACGTGCCAGCGGCATGACAACCCTCTCTGCAGAACTTTCCAGTCGAC
TCTCAACGGCACCCGACCTGACAGCGTTCAGCGACCCGCGCCAGTTCCCCGCGCTGC
CCTCCATCTCCGACCCCCGCATGCACTATCCAGGCGCCTTCACCTACTCCCCGACGCC
GGTCACCTCGGGCATCGGCATCGGCATGTCGGCCATGGGCTCGGCCACGCGCTACCA
CACCTACCTGCCGCCGCCCTACCCCGGCTCGTCGCAAGCGCAGGGAGGCCCGTTCCA
AGCCAGCTCGCCCTCCTACCACCTGTACTACGGCGCCTCGGCCGGCTCCTACCAGTTC
TCCATGGTGGGCGGCGAGCGCTCGCCGCCGCGCATCCTGCCGCCCTGCACCAACGCC
TCCACCGGCTCCGCGCTGCTCAACCCCAGCCTCCCGAACCAGAGCGACGTGGTGGAG
GCCGAGGGCAGCCACAGCAACTCCCCCACCAACATGGCGCCCTCCGCGCGCCTGGAG
GAGGCCGTGTGGAGGCCCTACGAGGTGCACACCAACCAGGACCCCCTGGACGCCGAA
GTCCATACAAATCAGGATCCTCTGGATGCCGAAGTGCACACCAATCAGGATCCCCTGGA
CGCTGGAAGCGGAGCTACTAACTTCTCTCTGTTAAAGCAAGCAGGAGACGTGGAAGAA
AACCCCGGTCCCATGGTGAGCAAGGGCGAGGAGCTGTTCACCGGGGTGGTGCCCATC
CTGGTCGAGCTGGACGGCGACGTAAACGGCCACAAGTTCAGCGTGTCCGGCGAGGGC
GAGGGCGATGCCACCTACGGCAAGCTGACCCTGAAGTTCATCTGCACCACCGGCAAGC
TGCCCGTGCCCTGGCCCACCCTCGTGACCACCCTGACCTACGGCGTGCAGTGCTTCA
GCCGCTACCCCGACCACATGAAGCAGCACGACTTCTTCAAGTCCGCCATGCCCGAAGG
CTACGTCCAGGAGCGCACCATCTTCTTCAAGGACGACGGCAACTACAAGACCCGCGCC

GAGGTGAAGTTCGAGGGCGACACCCTGGTGAACCGCATCGAGCTGAAGGGCATCGAC
TTCAAGGAGGACGGCAACATCCTGGGGCACAAGCTGGAGTACAACTACAACAGCCACA
ACGTCTATATCATGGCCGACAAGCAGAAGAACGGCATCAAGGTGAACTTCAAGATCCGC
CACAACATCGAGGACGGCAGCGTGCAGCTCGCCGACCACTACCAGCAGAACACCCCC
ATCGGCGACGGCCCCGTGCTGCTGCCCGACAACCACTACCTGAGCACCCAGTCCGCC
CTGAGCAAAGACCCCAACGAGAAGCGCGATCACATGGTCCTGCTGGAGTTCGTGACCG
CCGCCGGGATCACTCTCGGCATGGACGAGCTGTACAAGTGAGGATCCTGATCATAATCA
GCCATACCACATTTGTAGAGGTTTTACTTGCTTTAAAAAACCTCCCACACCTCCCCCTGA
ACCTGAAACATAAAATGAATGCAATTGTTGTTGTTAACTTGTTTATTGCAGCTTATAATGGT
TACAAATAAAGCAATAGCATCACAAATTTCACAAATAAAGCATTTTTTTCACTGCATTCTAG
TTGTGGTTTGTCCAAACTCATCAATGTATCTTAGGGTCAGCCCACACCACCCAGCCCCC
ACGCCCAACCCTCGTGCCTCCCTGAACCACTCCACTGCCTTTAACCCTCAGCCTCAGA
GTCAGATGCAGGGTAAGTACCAGATGGAGCCCACTGCCCGCCTCTCCTGCACCTGGGC
CACCACCCACAACTGGCCCCCATGTGCACACACCTTCCCAGACCAACTGGGGTTTCCC
CTTGATGCTCAGAGAAAGGCCTCGAACCAACAGCACCACCTGGGAGCTGTTGGAAATG
CAGAGTCTTGGGCCCCACCCACCCCAGACCCACCGAGTCAGATCTGCATTGTAACCAG
ATCCCCACAAGGAAAAGCACTGCTACAGAGGATACGAGGAGCTCTGGGTATGGATATCC
ATATTGGATAGATAGCTCTCTTGAATTTTTTAAGAGTTAGCTTTGGCTATGCTATTTTTTTAC
CTACCCATTAGGCTTGAAGACACACACGCAGACACACACACACACACACGTCAAGCAAA
GTGAAGATGGGATGTTTCAAGTTCTCGCTATTGCCAGATTATTTGTGGGTTTTGTATCTAG
TGTTTTTTTATTATTAAGAAATAGTTGAAATGTATGGATGTCATCACATCAAGGGTGTTATTT
GCAAATCAATAGAGAATGCAGGTCCCCCCAGCCCATGGGGCTAGCTGGCAATTACTAAA

806 GCGCTGTAAGATGCAATAATTGCCTAAGGCCCACTGTGCCAAATTAGATAATACAAGAAG

807 TTCATTTACACTGTAGACCAGTGACGTCAATGACTGTTTGCTCTGTGATACCGTTTCGTC

808 AAGATGTTTGCTTGCAAAGG

**Primer list:**

| Primer name | Sequence | Purpose |
|---|---|---|
| pc3.1-RUNX1b-F | TTGGTACCGAGCTCGGATCCGCCACCATGCGTATCCCCGTAGATGCCAGCACGAGCCGCCG | pcDNA3.1-RUNX1b |
| pc3.1-GFP-F | TTGGTACCGAGCTCGGATCCGCCACCATGGTGAGCAAGGGCGAGGAG | pcDNA3.1-EGFP-RUNX1b |
| pc3.1-GFP-RUNX1b-r | CGGCGGCTCGTGCTGGCATCTACGGGGATACGCATCTTGTACAGCTCGTCCATGCC | pcDNA3.1-EGFP-RUNX1b |
| pc3.1-GFP-RUNX1b-f | GGCATGGACGAGCTGTACAAGATGCGTATCCCCGTAGATGCCAGCACGAGCCGCCG | pcDNA3.1-EGFP-RUNX1b |
| pc3.1-mCherry-F | TTGGTACCGAGCTCGGATCCGCCACC ATGGTGAGCAAGGGCGAGGAG | pcDNA3.1-mCherry-RUNX1b |
| pc3.1-mCherry-RUNX1b-R | CGGCGGCTCGTGCTGGCATCTACGGGGATACGCATCTTGTACAGCTCGTCCATGCC | pcDNA3.1-mCherry-RUNX1b |
| pc3.1-mCherry-RUNX1b-F | GGCATGGACGAGCTGTACAAGATGCGTATCCCCGTAGATGCCAGCACGAGCCGCCG | pcDNA3.1-mCherry-RUNX1b |
| pc3.1-RUNX1b-R | ACGGGCCCTCTAGACTCGAGTCAGTAGGGCCTCCACACGG | pcDNA3.1-mCherry/EGFP-RUNX1b |
| pc3.1-CBFb-F | CGTTTAAACTTAAGCTTGGTACCATGCCGCGCGTCGTGCC | pcDNA3.1-CBFb |

| | | |
|---|---|---|
| pc3.1-CBFb-R | ACGGGCCCTCTAGACTCGAGTTAACGAAGTTTGAGGTCATCACCACC | pcDNA3.1-CBFb |
| attB-FLAG-RUNX1b-F | GATATCACCGCAAGAGCTCCACGCCACCATGGATTACAAGGATGACGACGATAAGGGCGATTACAAGGATGACGACGATAAGATGCGTATCCCCGTAGATGCCAGCACGAGCCGCCG | attB-RUNX1-IRES-mCherry |
| attB-RUNX1b-R | TCAGACCGGTGAATTCTTAAGTTATCAGTAGGGCCTCCACACGG | attB-RUNX1-IRES-mCherry |
| cl20c-2Flag-RUNX1b-F | TTCTCTAGGCGCCGGAATTCGCCACCATGGATTACAAGGATGACGACGATAAGGGCGATTACAAGGATGACGACGATAAGATGCGTATCCCCGTAGATGCCAGCACGAGCCGCCG | cl20c-MSCV-2Flag-RUNX1b-IRES-GFP |
| cl20c-RUNX1b-R | TGCATGGATCCCTAGGAATTCTCAGTAGGGCCTCCACACG | cl20c-MSCV-2Flag-RUNX1b-IRES-GFP |
| cl20c-MSCV-IRES-mCherry-F | CCCCCGAACCACGGGGACGTGGTTTTCCTTTGAAAAACACGATAATACCATGGTGAGCAAGGGCGAGGAGGA | cl20c-MSCV-IRES-mCherry |
| cl20c-MSCV-IRES-mCherry-R | TATACGGCATCGATGCGGCCGCTTCACTTGTACAGCTCGTCCA | cl20c-MSCV-IRES-mCherry |
| cl20c-MSCV- | TCCTTCTCTAGGCGCCGGAATTCGCCACCATG | cl20c-MSCV-JAK3- |

| | | |
|---|---|---|
| JAK3-IRES-mCherry-F | GCACCTCCAAGTGAAGA | IRES-mCherry |
| cl20c-MSCV-JAK3-IRES-mCherry-R | TGCATGGATCCCTAGGAATTCCTATGAAAAGGACAGGGAGTG | cl20c-MSCV-JAK3-IRES-mCherry |
| JAK3-M511I-mutagenesis-F | CCAATACCAGCTGAGTCAGATAACATTTCACAAGATCC | cl20c-MSCV-JAK3(M511I)-IRES-mCherry |
| JAK3-M511I-mutagenesis-R | GGATCTTGTGAAATGTTATCTGACTCAGCTGGTATTGG | cl20c-MSCV-JAK3(M511I)-IRES-mCherry |
| p.K117*-F | CTGCCCATCGCTTTCTAGGTGGTGGCCCTAG | Mutagenesis |
| p.K117*-R | CTAGGGCCACCACCTAGAAAGCGATGGGCAG | Mutagenesis |
| p.Q213fs-F | GCAGAAACTAGATGATAGACCAAGCCCGGGAGCTTG | Mutagenesis |
| p.Q213fs-R | CAAGCTCCCGGGCTTGGTCTATCATCTAGTTTCTGC | Mutagenesis |
| p.R232fs-F | CTGGAGCAGCTGCGCGCACAGCCATGAG | Mutagenesis |
| p.R232fs-R | CTCATGGCTGTGCGCGCAGCTGCTCCAG | Mutagenesis |
| p.N153Y-F | CTACCGCAGCCATGAAGTACCAGGTTGCAAG | Mutagenesis |
| p.N153Y-R | CTTGCAACCTGGTACTTCATGGCTGCGGTAG | Mutagenesis |

| | | |
|---|---|---|
| p.S141fs-F | CAATGATGAAAACTACTCAACCGGCTGAGCTG AGAAATG | Mutagenesis |
| p.S141fs-R | CATTTCTCAGCTCAGCCGGTTGAGTAGTTTTCA TCATTG | Mutagenesis |
| p.Y287*-F | CGATCAGTCCTACCAATAGCTGGGATCCATTG C | Mutagenesis |
| p.Y287*-R | GCAATGGATCCCAGCTATTGGTAGGACTGATC G | Mutagenesis |
| p.K110Q-F | GGCGCTGCAACCAGACCCTGCCCATC | Mutagenesis |
| p.K110Q-R | GATGGGCAGGGTCTGGTTGCAGCGCC | Mutagenesis |
| p.P275L-F | CAAGGCAGATCCAACTATCCCCACCGTGGTC | Mutagenesis |
| p.P275L-R | GACCACGGTGGGGATAGTTGGATCTGCCTTG | Mutagenesis |
| p.T246M-F | ACCCAGCCCCCATGCCCAACCCTCGT | Mutagenesis |
| p.T246M-R | ACGAGGGTTGGGCATGGGGGCTGGGT | Mutagenesis |
| p.313_317del-F | GCGGCATGACAACCCTTTCCAGTCGACTCTC | Mutagenesis |
| p.313_317del-R | GAGAGTCGACTGGAAAGGGTTGTCATGCCGC | Mutagenesis |
| p.M418V-F | CTACCAGTTCTCCGTGGTGGGCGGCGAG | Mutagenesis |
| p.M418V-R | CTCGCCGCCCACCACGGAGAACTGGTAG | Mutagenesis |
| p.G365R-F | CACCTCGGGCATCCGCATCGGCATGTC | Mutagenesis |
| p.G365R-R | GACATGCCGATGCGGATGCCCGAGGTG | Mutagenesis |
| p.P359R-F | TACTCCCCGACGCGGGTCACCTCG | Mutagenesis |

| | | |
|---|---|---|
| p.P359R-R | CGAGGTGACCCGCGTCGGGGAGTA | Mutagenesis |
| p.S318_S319delinsX-F | CTCTGCAGAACTTTAGCTCCAGTCGACTCTCAAC | Mutagenesis |
| p.S318_S319delinsX-R | GTTGAGAGTCGACTGGAGCTAAAGTTCTGCAGAG | Mutagenesis |
| 365-OutArm-F | CCTGGGCGGTAAATTCTGATAG | CHASE-KI validation |
| 365-LHA-F2 | GGCATAGCATCATGGGTAGTC | CHASE-KI validation |
| 232-OutArm-F2 | CAGGACTGGCTCTGGTTTAAG | CHASE-KI validation |
| 232-LHA-F2 | CCCAAATTCAGCTGGCATATC | CHASE-KI validation |
| 287-OutArm-F2 | CATTTGCCTAAGAATAGCGTTGG | CHASE-KI validation |
| 287-LHA-F2 | GGCAGATACTTGGACTTGAGTAG | CHASE-KI validation |
| mCherry-nest-R3 | CGCAGCTTCACCTTGTAGAT | CHASE-KI validation |
| EGFP-nest-R | AGACGTTGTGGCTGTTGTAG | CHASE-KI validation |

| | | |
|---|---|---|
| RUNX1-Stop-gRNA+PAM | GGAGGCCCTACTGAGGCGCCAGG | CHASE-KI validation |
| RUNX1-Q232fs-gRNA40+PAM | TCAGTGAACTGGAGCAGCTGCGG | CHASE-KI validation |
| RUNX1-p.Y287*-gRNA76+PAM | AGGCAATGGATCCCAGGTATTGG | CHASE-KI validation |
| Seq-HA | TAGGATCCTGCATAGTCCGG | CHASE-KI validation |
| Seq-TY1 | GTGTGCACTTCGGCATCCA | CHASE-KI validation |
| Seq-CDS | CCACCATGGAGAACTGGTAG | CHASE-KI validation |
